# Large language models and retrieval augmented generation for complex clinical codelists: evaluating performance and assessing failure modes

**DOI:** 10.64898/2026.04.23.26351098

**Authors:** Julian Matthewman, Spiros Denaxas, Sinéad M Langan, Jeffery Painter, Andrew Bate

## Abstract

**Objectives:** Large language models (LLMs) have shown promise in creating clinical codelists for research purposes, a time-consuming task requiring expert domain knowledge. Here, we evaluate the performance and assess failure modes of a retrieval augmented generation (RAG) approach to creating clinical codelists for the large and complex medical terminology used by the Clinical Practice Research Datalink (CPRD).

**Materials & Methods:** We set up a RAG system using a database of word embeddings of the medical terminology that we created using a general-purpose word embedding model (gemini-embedding). We developed 7 reference codelists presenting different challenges and tagged required and optional codes. We ran 168 evaluations (7 codelists, 2 different database subsets, 4 models, 3 epochs each). Scoring was based on the omission of required codes, and inclusion of irrelevant codes. We used model-grading (i.e., grading by another LLM with the reference codelists provided as context) to evaluate the output codelists (a score of 0% being all incorrect and 100% being all correct).

**Results:** We saw varying accuracy across models and codelists, with Gemini 3 Pro (Score 43%) generally performing better than Claude Sonnet 4.6 (36%), Gemini 3 Flash, and OpenAI GPT 5.2 performing worst (14%). Models performed better with shorter target codelists (e.g., Eosinophilic esophagitis with four codes, and Hidradenitis suppurativa with 14 codes). For example, all models consistently failed to produce a complete Wrist fracture codelist (with 214 required codes). We further present evaluation summaries, and failure mode evaluations produced by parsing LLM chat logs.

**Discussion:** Besides demonstrating that a single-shot RAG approach is currently not suitable for codelist generation, we demonstrate failure modes including hallucinations, retrieval failures and generation failures where retrieved codes are not used.

**Conclusions:** Our findings suggest that while RAG systems using current frontier LLMs may create correct clinical codelists in some cases, they still struggle with large and complex terminologies and codelists with a large number of codes. The failure mode we highlight can inform the creation of future workflows to avoid failures.

## Background

Codelists are required to extract information from routinely collected health data such as electronic health records and their correctness is important to ensure trustworthy results when using these data for research. Developing codelists is an iterative multi-step process requiring clinical domain knowledge.(1)

Codelists necessarily vary by clinical concept, but also by the nature of the healthcare system that is being used and the recording rules of any given clinical support system and/or healthcare database. Therefore, new or adapted code lists need to be developed frequently when epidemiological analyses are planned, with several tools and frameworks available.(2–4) There is a need to minimize accidental variability between codelists, ensure they can be readily re-used and are accessible, but also that when new code lists are needed they can be developed quickly and with high quality.

Large language models, which are now readily available, encode clinical knowledge.(5) While the use of LLMs has already shown promise in creating phenotyping algorithms with ICD-10 codes,(6,7) other terminologies are more complex. For example, while a codelist for asthma in ICD-10 may only contain the codes “J45 Asthma” and “J46 Status asthmaticus” (and all descendant codes), in SNOMED or Read it may contain hundreds of codes, including for diagnoses, symptoms, administration records, referrals, and treatments that are not always structured hierarchically and may be updated frequently. Therefore, there is a need to evaluate the use of LLMs in scenarios where the terminology is complex or new.

There is also a need to assess the types of mistakes LLMs can make while generating codelists (“failure modes”). For example, LLMs can hallucinate (return a code that does not exist in the target terminology). Retrieval augmented generation (RAG) may partially address the issue of hallucinations.(8) Instead of relying on information on codes that is encoded in LLMs training data, codes from a knowledge store can be retrieved in an initial step and provided as context to the LLM in the generation of its response. In RAG workflows, the knowledge store contains word embeddings, i.e., numerical representations of words, which may also help find synonyms of a given concept.

However, RAG systems may still miss required codes or include irrelevant codes, for example by failing to retrieve a code from the knowledge store, failing to use a retrieved code or hallucinating despite retrieval.

Our objective was to evaluate the suitability of LLMs in the complex but common scenario where new codelists need to be created for a specific medical terminology. To do this, we scored performance of a RAG system in the creation of codelists and assessed failure modes.

## Methods

### Terminology

We used the September 2025 build of the medical code browser file of the Clinical Practice Research Datalink (CPRD) which contains medical codes available to GP staff and contains a combination of SNOMED-CT, Read and local EMIS codes.(9) SNOMED CT (Systemized Nomenclature of Medicine – Clinical Terms) is a comprehensive and internationally validated system to record clinical information that superseded the use of Read codes in UK primary care.(10) Local EMIS codes are codes specific to the EMIS general practice software. The entire medical code browser is mapped to SNOMED CT, while only a subset of terms is mapped to Read codes. No patient data were used.

### Reference codelist development

We developed 7 reference codelists. In the development of codelists, judgement calls need to be made, often based on limited evidence, e.g., with which certainty a code needs to indicate that a person may currently have a disease. Therefore, we tagged codes as required and optional.(1) Codelists were developed iteratively and tested with the RAG system until any mistakes by the RAG system were unambiguous mistakes of the LLM, rather than human error or omission in the reference codelist creation (this was done by iteratively checking the evaluation logs). The development of the reference codelists was informed by our previous experience in developing codelists for the CPRD database and our clinical experience.

### Creating word embeddings

We created a word embedding knowledge store of the terms contained in the CPRD Aurum September 2025 Medical Browser using the gemini-embedding model. We then added the respective codes to the terms and their embeddings so that they could be retrieved by the LLM, i.e., the embedding was only for the term, e.g., “Psoriasis”, and not the code, e.g., “M161z”.

### Setup of Retrieval Augmented Generation System

We set up a retrieval augmented generation workflow using the `ellmer`(11)(for making requests to LLMs within an R environment), and `ragnar`(12) (for implementing RAG workflows) R packages. We registered a tool that allowed LLMs to retrieve the 100 most semantically similar terms from the word embeddings. Multiple tool calls could be performed during each codelist generation. We provided the following system prompt: *“Create a codelist for the given clinical condition in the input. Include all codes that when recorded in a person’s medical record would indicate that the person currently has the condition. Use only codes from the knowledge store*.*”*

### Evaluation

We used the `vitals`(13) R package for evaluation. We used “model grading” (also known as “LLM-as-a-judge”, i.e., each LLM output was evaluated by another LLM that had the gold standard codelist provided as context. Model grading allowed us to evaluate many hundreds of LLM produced codelists without needing to specify a structured output type. The following scoring instruction were used: *“In the target codelist in the ‘required’ column, 1 denotes a required code, 0 denotes an optional code. Score as correct only if all required codes are included and no codes not in the target codelist (either marked ‘required’ or ‘optional’) are included in the final codelist. Score as partially correct if all required codes are included but there are also codes included that are not in the target codelists. Otherwise score as incorrect*.*”* We performed each evaluation 3 times (across 3 “epochs”) and for the Gemini 3 Pro (preview), Gemini 3 Flash (preview), OpenAI GPT 5.2, and the Anthropic Claude Sonnet 4.6 models. A score was calculated as the proportion of correct responses out of all responses for all codelists and epochs, with partially correct responses counting half of a correct response. Evaluation summaries were drafted with Gemini 3 Pro and edited and checked for correctness.

### Failure modes

To check for failure modes, we converted the LLM responses into structured data (using Claude Sonnet 4.6). Firstly, we checked for codes in the generated codelists that do not exist in the original medical terminology (“hallucinations”). Secondly, we checked whether errors were due to failure at the retrieval or the generation step by assessing the presence of required codes in the retrieval logs and assessing if any required retrieved codes were not used in the generation step.

### Alternative workflows

We performed the following alternative workflows: 1. Used an alternative retrieval method using string searching, 2. Changed the number of retrieved codes per tool call from 100 to 200, 3. Used a biomedical embeddings model (instead of a general-purpose model) to create the knowledge store, 4. Used agentic coding to create codelists.

### Workflow with alternative retrieval method: string searching

In our main workflow terms were retrieved based on their semantic similarity from a knowledge store created with an embeddings model. As an alternative method we set up a workflow where terms that matched a searchterm were retrieved directly from the medical terminology. This had the advantage of not needing to specify a cut-off for the number of most semantically similar codes to retrieve, however the disadvantage of only retrieving exact string matches (e.g., if the LLM searched for “asthma” it would retrieve “asthma” and “asthmatic”, but never related terms not containing “asthma” such as “inhaler”). We registered two tools with the LLMs, firstly a tool that did a case-insensitive and position-insensitive (i.e., matching words in any order) search of the term column, and secondly, a tool that performed a search of the code column. Due to this workflow making more API request, we could not perform this evaluation with Gemini 3 Pro as the workflow exceeded the daily request limit.

### Alternative embeddings model

In our main workflow we used a general-purpose word embedding model (gemini-embedding). As an alternative embeddings model, we used the MedCPT-query model, a model trained on user click logs from PubMed.(14)

### Agentic coding

In our main workflow, codelists were generated directly by the LLM. As an alternative method we trialled a workflow using agentic coding with Claude Code, where the LLM agent would create an R script that would create the codelist.

## Data availability

All evaluations are included in the study repository at https://github.com/julianmatthewman/llmcodelists_public and can be browsed with an evaluation viewer at https://julianmatthewman.github.io/llmcodelists_public/.

## Results

We saw varying accuracy across models and codelists, with [Gemini 3 Pro (Score 43%)](https://julianmatthewman.github.io/llmcodelists_public/#/logs/2026-03-04T11-47-58%2B00-00_RAG_5ab6567c11a9be5c54356c.json) generally performing better than [Claude Sonnet 4.6 (36%)](https://julianmatthewman.github.io/llmcodelists_public/#/logs/2026-02-19T12-55-50%2B00-00_RAG_5ab6567c11a9be5c54356c.json), and [Gemini 3 Flash (19%)](https://julianmatthewman.github.io/llmcodelists_public/#/logs/2026-02-04T12-03-25%2B00-00_RAG_f5178a12554e983adb7209.json), with [OpenAI GPT 5.2 performing worst (14%)](https://julianmatthewman.github.io/llmcodelists_public/#/logs/2026-01-21T12-26-16%2B00-00_RAG_f5178a12554e983adb7209.json). Models performed better with shorter target codelists (e.g., Eosinophilic esophagitis with 3 required codes, and Hidradenitis suppurativa with 8 required codes). For example, all models consistently failed to produce complete wrist fracture (with 214 required codes) and myocardial infarction (with 132 required codes) codelists. Claude Sonnet 4.6 was the only model to produce codelists that were scored as partially correct, i.e., including all required codes but also irrelevant codes.

When subsetting the medical terminology to only include Read codes (which reduced target codelist sizes substantially), performance across all models was better ([Gemini 3 Pro: 81%; Claude Sonnet 4.6: 60%; Gemini 3 Flash: 69%; OpenAI GPT 5.2: 29%](https://julianmatthewman.github.io/llmcodelists_public/#/logs/2026-01-26T10-41-03%2B00-00_RAG_f5178a12554e983adb7209.json)), Gemini 3 Pro only failing to create the [Wrist Fracture codelist](https://julianmatthewman.github.io/llmcodelists_public/#/logs/2026-01-26T10-41-03%2B00-00_RAG_f5178a12554e983adb7209.json/samples/sample/7/1/) (160 required codes). Figure 2 shows scores, and Table 2 and Table 3 contain summaries of missed and irrelevant codes for the full evaluation and the evaluation with Read codes only, respectively.

**Table 1:** Examples of required, optional, and irrelevant codes for each codelist.

| Codelist | Required | Optional | Irrelevant |
| --- | --- | --- | --- |
| All codelists | Diagnosis codes indicating that the person currently has the disease. | For chronic conditions, (personal) medical history codes. Administrative codes. | Family history codes |
| Eosinophilic oesophagitis | J1017 Eosinophilic oesophagitis, ^ESCTEO508265 Eosinophilic esophagitis, and ^ESCT1165474 Food-induced eosinophilic oesophagitis. | ^ESCTEO508286 Eosinophilic ulcer of oesophagus. | Other oesophagitis, general eosinophilia. |
| Hidradenitis suppurativa | Hidradenitis suppurativa and its synonym Verneuil's disease. | Codes specifying "Hidradenitis" without "suppurativa", Follicular occlusion triad/tetrad (Hidradenitis suppurativa is one of the diseases of these syndromes). | Neutrophilic eccrine hidradenitis (a complication of chemotherapy). |
| Atopic Dermatitis | Dermatitis or eczema codes specifically mentioning "atopic" or "intrinsic". Flexural and infantile eczema as these are most commonly atopic. Besnier's prurigo (an older term for atopic dermatitis). | More unspecific forms of eczema that could be atopic, neurodermatitis, constitutional eczema (an older term with unclear usage). | Non-atopic forms of eczema such as contact eczema, seborrheic eczema, stasis eczema, varicose eczema, psoriasis. |
| Vascular dementia | Vascular, atherosclerotic, infarct dementias, Binswanger's disease (a form of small-vessel vascular dementia). | Mixed Alzheimer's and vascular dementias, cerebrovascular disease, sequelae of vascular dementia. | Alzheimer's dementia, confusional states of cerebrovascular origin. |
| Psoriasis | All forms of psoriasis, psoriatic nail involvement. | Psoriatic arthritis, Seborrheic psoriasis, Köbner psoriasis, PASI - psoriasis area and severity index, "Psoriasis and similar disorders". | Psoriasiform conditions, Arthritis mutilans (occurs mainly in people who have pre-existing psoriatic arthritis but can occur in rheumatoid arthritis). |
| Myocardial infarction | Infarctions specifying "myocardial" or a location in the heart (e.g., anteroapical infarction), or and ECG (e.g., acute non-Q wave infarction) | Subsequent, recurrent, postoperative or procedural myocardial infarctions. Complications, syndromes. Vascular occlusions and thrombosis without specifying an infarct. | Old infarctions and personal history of infarction. Infarcts elsewhere (e.g., spinal infarct). |
| Wrist fracture | Fractures of the carpal bones and radiocarpal joint, distal radius fractures, and Colles, Smith's, Barton's, Chauffeurs, and Hutchinson's fractures | Fractures of the radioulnar joint, distal parts of forearm bones, proximal parts of metacarpus or thumb, sesamoid bones (could be pisiform of the wrist but also other sesamoid bone in hand). Surgeries and reductions. Codes using "#" to mean fractures (e.g., #Carpal bones). | Non-proximal or unspecified fractures of the metacarpal bones, non-distal fractures of radius or ulna, fractures elsewhere (e.g., foot). |

**Table 2:** Full evaluation - Summary of omissions of required codes and inclusions of irrelevant codes.

| codelist | Omissions of required codes | Inclusion of irrelevant codes and hallucinated codes |
| --- | --- | --- |
| **Eosinophilic esophagitis** (3 required, 1 optional) | Claude Sonnet 4.6, Gemini 3 Pro, and GPT 5.2 correctly identified all required codes across all epochs. Gemini 3 Flash missed all required codes in Epoch 1: J1017 **Eosinophilic oesophagitis**, ^ESCT1165474 **Food-induced eosinophilic oesophagitis**, and ^ESCTEO508265 **Eosinophilic esophagitis**. | No irrelevant codes were included by any model across all evaluated epochs. Differences between models were limited to the inclusion of the optional code ^ESCTEO508286 **Eosinophilic ulcer of oesophagus**, which was provided by Claude Sonnet 4.6 and Gemini 3 Flash in certain runs. |
| **Hidradenitis suppurativa** (8 required, 6 optional) | GPT 5.2 consistently missed ^ESCTVE346453 **Verneuil's disease** across all three epochs. This same code was also omitted by Claude Sonnet 4.6 in Epoch 3 and Gemini 3 Flash in Epoch 2. Gemini 3 Pro was the most robust model, successfully identifying all required codes in every run. No other required codes were reported as missing across the submissions. | Claude Sonnet 4.6 included several irrelevant codes across epochs, such as ^ESCTDI375766 **Dissecting cellulitis of scalp**, Myu6J **Disorder of apocrine gland**, and Myu6D **Other apocrine sweat disorders**. In contrast, Gemini 3 Flash, Gemini 3 Pro, and GPT 5.2 restricted their inclusions to codes listed in the target criterion. |
| **Atopic eczema** (56 required, 155 optional) | Claude Sonnet 4.6 included all required codes across all evaluated epochs. Gemini 3 Pro had variable performance, with one complete submission (Epoch 3) but missing M115 **Besnier's prurigo** and ^ESCTXE666127 **Xerosis due to atopic dermatitis** in Epoch 1, and failing to identify various specific forms like ^ESCTDI512288 **Discoid atopic dermatitis** and ^ESCTPH512297 **Photosensitive atopic dermatitis** in Epoch 2. Gemini 3 Flash and GPT 5.2 frequently omitted M115 **Besnier's prurigo** and ^ESCTFO512293 **Follicular atopic dermatitis**. GPT 5.2 had the most significant omissions, regularly failing to include Read codes like M113 **Flexural eczema** and M114 **Allergic (intrinsic) eczema**. | Claude Sonnet 4.6 hallucinated codes such as ^ESCTAT668133 **Atopic dermatitis of eyelid**, ^ESCTAC598664 **Eczema of leg**, ^ESCTPS512296 **Psoriasisiform atopic eczema (psoriasisiform subtype)**. It was also the only model to include codes entirely outside the target list, such as ^ESCTNO696959 **Non-IgE-mediated atopic disorder**, ^ESCTAT697957 **Atopic IgE-mediated allergic disorder**, and ^ESCTAT668096 **Atopic keratoconjunctivitis**. In contrast, Gemini 3 Flash, Gemini 3 Pro, and GPT 5.2 did not include any irrelevant codes, though they varied in their inclusion of optional codes from the target list or omission of required ones. |
| **Vascular dementia** (63 required, 41 optional) | Claude Sonnet 4.6 initially missed F21y2 **Binswanger's disease** but achieved full coverage by the third epoch. Gemini 3 Flash consistently failed to include types like E0042 **Arteriosclerotic dementia with paranoia** and E0043 **Arteriosclerotic dementia with depression**. Gemini 3 Pro performed best, missing only E004-1 **Multi-infarct dementia** in the first epoch and capturing all required codes thereafter. GPT 5.2 performed poorly across all attempts, missing a wide array of required codes such as F21y2 **Binswanger's disease**, E0041 **Arteriosclerotic dementia with delirium**, and ^ESCTSU396465 **Subcortical arteriosclerotic encephalopathy**. | Claude Sonnet 4.6 hallucinated **F5074 Binswanger's disease**, ^ESCT1472649 **Behavioural disturbance due to multi-infarct dementia** and **F26227 Cerebral degeneration due to cerebrovascular disease**. It also included ^ESCTDE751279 **Dementia associated with cerebral anoxia**. Gemini 3 Pro included several irrelevant Alzheimer-mixed codes in its third epoch, such as Eu002 [X]**Dementia in Alzheimer's dis, atypical or mixed type** and EMISICD10[F0020 **Dementia in Alzheimer's dis, atypical or mixed type, without additional symptoms**. Gemini 3 Flash and GPT 5.2 did not include any irrelevant codes. |
| **Psoriasis** (76 required, 36 optional) | Claude Sonnet 4.6 was the most reliable, missing required codes only in its third epoch, such as Nyu13 **Psoriasis with arthropathy**, and ESCTFL1 **Flexural psoriasis**. Gemini 3 Pro exhibited a highly consistent error pattern, failing to include ^ESCTPS512393 **Psoriatic nail involvement** and ^ESCTPS512395 **Psoriatic onycholysis** across all three runs. Gemini 3 Flash missed numerous codes such as ^ESCTLO382192 **Localised pustular psoriasis**. GPT 5.2 frequently omitted M161H **Erythrodermic psoriasis**, and M161J **Flexural psoriasis**. Both Gemini 3 Flash and GPT 5.2 also shared omissions such as M166 **Palmoplantar pustular psoriasis** and ^ESCTPS512392 **Psoriatic nail dystrophy**, and M1612 **Psoriasis circinate**. | Claude Sonnet 4.6 consistently included non-target codes such as M143 **Impetigo herpetiformis**, M161G **Acrodermatitis continua**, ^ESCTAC666403 **Acrodermatitis continua of Hallopeau**, and EMISNQA160 **Palmoplantar pustulosis**. Gemini 3 Flash uniquely introduced the M162 series related to **Parapsoriasis**. GPT 5.2 hallucinated codes, specifically ^ESCT142366 **Acute palmoplantar pustular psoriasis**, and ^ESCT149855 **Chronic palmoplantar pustular psoriasis**. Gemini 3 Pro did not report any irrelevant codes in its submissions. |
| **Myocardial infarction** (132 required, 96 optional) | Across evaluations, models consistently failed to include required anatomical and clinical subtypes. Claude Sonnet 4.6 missed specific site descriptors such as ^ESCTAC505627 **Acute Q wave infarction - anteroseptal**, and G36 **Myocardial infarction with complication**. Gemini 3 Flash frequently omitted broad clinical terms and CTV3 anatomical codes like G30-4 **Heart attack**, G301 **Acute anterior myocardial infarction**, and G303 **Acute inferoposterior infarction**. Gemini 3 Pro struggled with specialized pathology and transmural codes including ^ESCT1413295 **MINOCA - myocardial infarction with non-obstructive coronary artery**, and ^ESCTNS665139 **NSTEMI - Non-ST segment elevation MI**. GPT 5.2 recurrently missed core descriptors and ST-elevation classifications like G30-4 **Heart attack**, ^ESCTST665122 **STEMI - ST elevation myocardial infarction**, and ^ESCTNS665139 **NSTEMI - Non-ST segment elevation MI**. | Claude Sonnet 4.6 included historical and non-cardiac codes such as G32 **Old myocardial infarction**, G32-2 **Personal history of myocardial infarction**, and ^ESCTAC352556 **Acute arterial infarction of spinal cord**, and hallucinated codes like ^ESCTAC477597 **Acute myocardial infarction of atrium**. Gemini 3 Pro added administrative and vague codes including ^ESCTAC340067 **Acute infarct** and hallucinated **100361 Reason for care : Myocardial infarction (MI)**. GPT 5.2 mistakenly included non-specific ischaemic heart disease and history terms such as G31-99 **Acute/subacute IHD NOS**, G34-99 **Chr. ischaemic heart dis. NOS**, and 14AT **History of myocardial infarction**. Gemini 3 Flash was primarily noted for missing required codes rather than including irrelevant ones. |
| **Wrist fracture** (214 required, 125 optional) | Claude Sonnet 4.6 omitted detailed carpal bone codes such as S2402 **Closed fracture lunate**, and S2403 **Closed fracture triquetral**, alongside the S4C range including S4C0 **Closed fracture dislocation of wrist**. Gemini 3 Flash consistently missed distal radius subtypes like S234D **Closed extraarticular fracture of distal radius** and S2351 **Open Colles' fracture**. Gemini 3 Pro lacked codes for the lower end of the radius and ulna, specifically S23B **Fracture of lower end of radius** and S23C **Fracture of lower end of both ulna and radius**, as well as Syu63 [X]**Fracture of other carpal bone(s)**. GPT 5.2 frequently failed to provide named fractures such as S2347 **Closed Smith's fracture** and S2349 **Closed volar Barton's fracture**, and various open equivalents like S2351-1 **Smith's fracture - open**. | Claude Sonnet 4.6 included fractures outside the wrist region, such as S239 **Fracture of shaft of radius**, S2306 **Closed fracture radius, head**, and ^ESCTFR546196 **Fracture of radial head (Elbow)**. Gemini 3 Flash and Gemini 3 Pro both included ^ESCTTO651725 **Torus fracture of radius**, with Gemini 3 Pro further including unrelated injuries like ^ESCTPI546251 **Pilon fracture** and ^ESCTTR435831 **Trimalleolar fracture**. GPT 5.2 included generic radius fractures like ^ESCT1527958 **Fracture of right radius** and highly irrelevant spinal procedures including ^ESCT1503536 **Open reduction of fracture of bone of spine** and ^ESCT1491835 **Fixation of fracture of bone of spine**. |

**Table 3:** Evaluation of Read codes only - Summary of omissions of required codes and inclusions of irrelevant codes.

| codelist | Omissions of required codes | Inclusion of irrelevant codes |
| --- | --- | --- |
| **Eosinophilic esophagitis** (1 required, 0 optional) | All models (Claude Sonnet 4.6, Gemini 3 Flash, Gemini 3 Pro, and GPT 5.2) successfully identified the single required code J1017 **Eosinophilic oesophagitis** across all epochs. No codes were missed in any submission. | All models, including Claude Sonnet 4.6, Gemini 3 Flash, Gemini 3 Pro, and GPT 5.2, performed identically across all three epochs, providing only the required code and including no irrelevant codes in their submissions. |
| **Hidradenitis suppurativa** (1 required, 0 optional) | No required codes were missed. All models (Claude Sonnet 4.6, Gemini 3 Flash, Gemini 3 Pro, and GPT 5.2) correctly included the required code M25y1-1 **Hidradenitis suppurativa** across all epochs. | Claude Sonnet 4.6 and Gemini 3 Flash included irrelevant codes, while Gemini 3 Pro and GPT 5.2 were correct. Claude Sonnet 4.6 included M2yA **Acne conglobata**, M2yB **Acne fulminans**, M25 **Diseases of sweat glands**, and M2037 **Fox-Fordyce disease**. Gemini 3 Flash included unrelated terms like M06 **Other rheumatoid arthritis**, M2611 **Cellulitis and abscess of face**, and M0031 **Staphylococcal arthritis and polyarthritis**. Both models also hallucinated **M2440 Hidradenitis suppurativa**, **L73.2 Hidradenitis suppurativa**, and **Myu6D [V]Hidradenitis suppurativa**. |
| **Atopic eczema** (10 required, 28 optional) | Claude Sonnet 4.6, Gemini 3 Flash, and Gemini 3 Pro successfully identified all required codes across all epochs. In contrast, GPT 5.2 consistently missed M115 **Besnier's prurigo** in every epoch. | Claude Sonnet 4.6 included the most extraneous codes, notably systemic syndromes like C3912 **Wiskott-Aldrich syndrome** and C397 **Hyperimmunoglobulin E syndrome**. Gemini 3 Flash also introduced codes absent from the target list, specifically SN533 **Atopy**, A540-1 **Kaposi's varicelliform eruption**, and hallucinated **M11z1-99 Chronic Eczema**. In contrast, Gemini 3 Pro and GPT 5.2 were more precise, only including codes defined in the criterion. |
| **Vascular dementia** (16 required, 19 optional) | Claude Sonnet 4.6, Gemini 3 Flash, and Gemini 3 Pro included all required codes across all epochs. **GPT 5.2** consistently missed F21y2-1 **Binswanger's encephalopathy** in every epoch; in the third epoch, it additionally missed F21y2 **Binswanger's disease**. | Claude Sonnet 4.6 consistently included the irrelevant code G678 **Cerebral autosomal dominant arteriopathy with subcortical infarcts and leukoencephalopathy** (also referred to as **CADASIL**). Gemini 3 Flash similarly included G678 **Cerebral autosomal dominant arteriopathy with subcortical infarcts and leukoencephalopathy** in its third epoch alongside other extra codes. Gemini 3 Pro and GPT 5.2 did not include any irrelevant codes across their submissions. |
| **Psoriasis** (25 required, 10 optional) | Gemini 3 Flash, Gemini 3 Pro and Claude Sonnet 4.6 successfully identified all required codes. GPT 5.2 performed poorly, consistently missing required codes such as M161J **Flexural psoriasis** and Nyu13 **Psoriasis with arthropathy** (both missed in multiple epochs), as well as M161 **Other psoriasis**, and M166 **Palmoplantar pustular psoriasis** in individual epochs. | Claude Sonnet 4.6 frequently included irrelevant codes such as M161G **Acrodermatitis continua**, M143 **Impetigo herpetiformis**, and M1602 **Arthritis mutilans**. Gemini 3 Pro also included M161G **Acrodermatitis continua** in its second epoch. Gemini 3 Flash maintained strict adherence to the target list, while GPT 5.2 included several extra codes in its third epoch without associated terms. |
| **Myocardial infarction** (30 required, 41 optional) | Claude Sonnet 4.6, Gemini 3 Flash, and Gemini 3 Pro successfully included all required codes across all epochs. GPT 5.2 was the only model to miss required codes including G30-4 **Heart attack**, G30-99 **Myocardial Infarction**, **Acute ST segment elevation myocardial infarction** and several others. | Claude Sonnet 4.6 included acute complications such as G501 **Post infarction pericarditis** and G33z5 **Post infarct angina**. Gemini 3 Flash introduced broader diagnostic terms like G3115 **Acute coronary syndrome**. Gemini 3 Pro and GPT 5.2 both frequently included historical or healed condition codes, specifically G32 **Old myocardial infarction**, G32-1 **Healed myocardial infarction**, and various history markers such as 14AT **History of myocardial infarction** and hallucinated terms like **G32-99 Acute/subacute IHD NOS** and **G34y? Silent myocardial infarction**. |
| **Wrist fracture** (111 required, 49 optional) | Claude Sonnet 4.6 performed best, achieving one complete submission. When incomplete, it primarily missed specific synonym codes like S2341-98 **Radius/ulna-lower end-colles** and S2341-99 **Colles fracture**. Gemini 3 Flash consistently missed a broad range of required codes across epochs such as S2351 **Open Colles' fracture**, S235E **Open intra-articular fracture of distal radius**, and S2411 **Open fracture of the scaphoid**. Gemini 3 Pro frequently missed open variants of eponymous fractures, including S2351 **Open Colles' fracture** and S2357 **Open Smith's fracture**, alongside carpal codes like S2406 **Closed fracture trapezoid**. One epoch uniquely missed broad generic codes such as S242 **Fracture at wrist and hand level** and Syu65 **Fracture at wrist and/or hand level**. GPT 5.2 failed to identify fundamental base codes that were often found by other models, consistently missing S2341 **Closed Colles' fracture** and S2351 **Open Colles' fracture** across multiple epochs. It also repeatedly missed standard carpal codes like S2401 **Closed fracture of the scaphoid** and S2408 **Closed fracture hamate**. | Claude Sonnet 4.6 included numerous elbow and forearm shaft codes such as S2301 **Closed fracture olecranon**, S239 **Fracture of shaft of radius**, and S2303 **Closed Monteggia's fracture**, alongside generic forearm injury codes like S4 **Injury of upper limb** and S23 **Fracture of forearm**. It also included superior radio-ulnar joint codes like S4B21 **Closed fracture-subluxation superior radio-ulnar joint**. Gemini 3 Pro uniquely included S2342-1 **Dupuytren's fracture, radius - closed** and S2352-1 **Dupuytren's fracture, radius - open**. GPT 5.2 included generic radius codes such as S23x1 **Closed fracture of radius**. Gemini 3 Flash did not report any irrelevant codes. |

**Figure 1:**
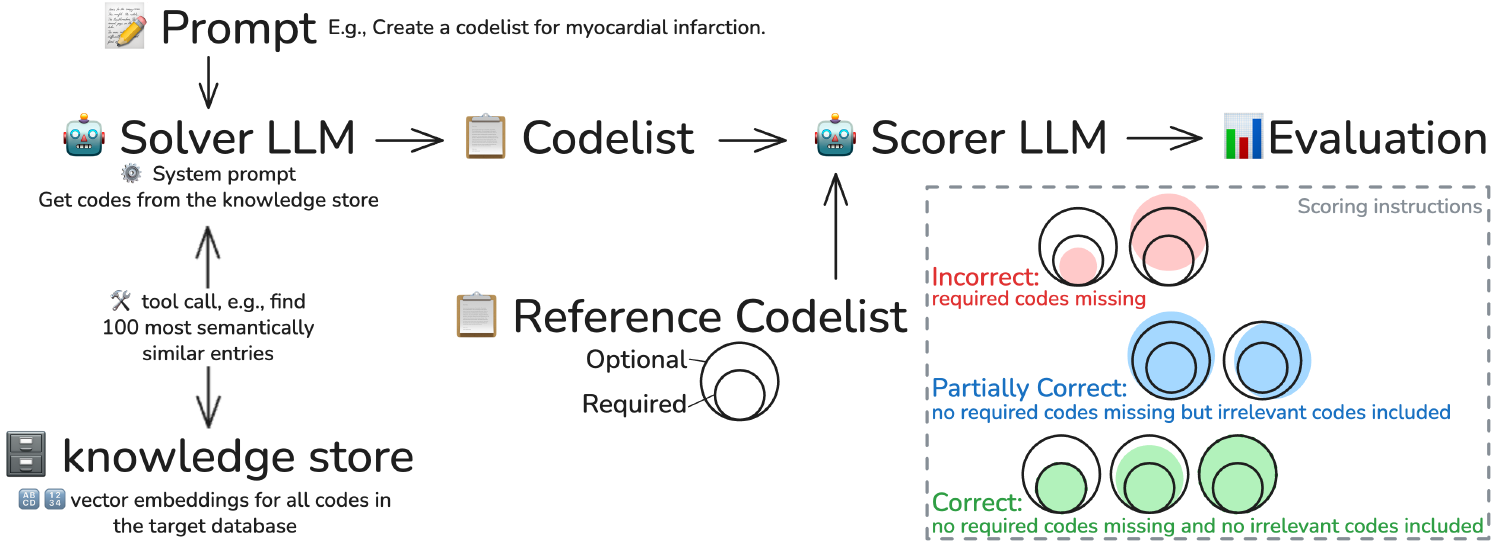
Diagram of codelist generation and evaluation workflows

**Figure 2:**
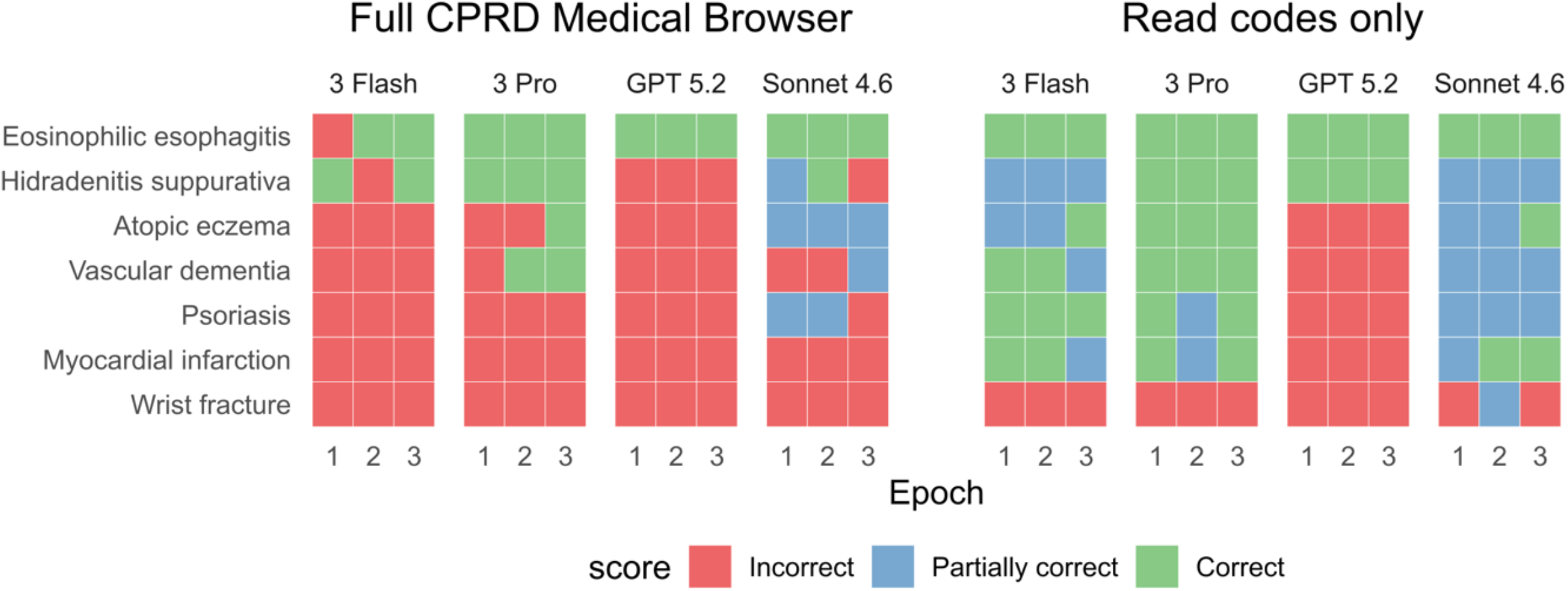
Evaluation results.

**Figure 3:**
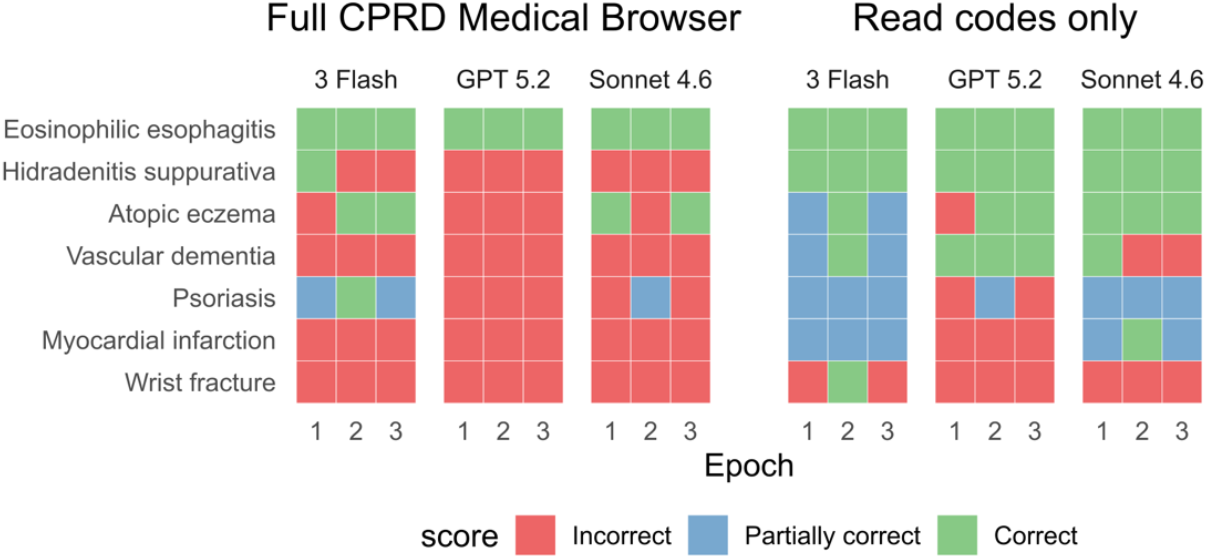
Evaluation results (string search workflow)

### Failure modes

We specifically examined three distinct failure modes: 1. hallucinations, i.e., the LLM generated codes that do not exist in the original code browser, 2. retrieval failures, i.e., the LLM did not retrieve required codes in the retrieval stage, and 3. generation failures, i.e., the LLM did not use required codes that were retrieved in the retrieval stage. This was possible by parsing the LLM chat logs which contain all tokens passed to and from the LLM including during tool calls. Exact string matches were used to check for retrieval and hallucinations, hence codelists with formatting differences (e.g., missing the “^” at the start of codes) flagged in the assessment of failure modes were still scored as correct in the main evaluation since the model-graded evaluation did not count formatting differences as mistakes.

Gemini 3 Pro and GPT 5.2 generally had the lowest proportion of epochs containing hallucinations (both 29%), followed by Gemini 3 Flash (43%) with Claude Sonnet 4.6 having the highest (52%) (also reflected in the larger proportion of partially correct scores for Claude Sonnet 4.6). The lists of hallucinated codes revealed that the number of hallucinated per epoch was also higher for Claude Sonnet 4.6 than for other models (Table 4, Supplementary Table 1).

**Table 4:** Summary of failure modes. Columns under the “Retrieval” spanner indicate the number of required codes from the reference codelist, the number of retrieved required codes, the number of not retrieved required codes and the number of required codes that were retrieved but unused. Columns under the “Hallucinations” spanner indicate the total number of codes returned, the number and percentage of codes hallucinated (i.e., these did not exist in the original medical code browser). Exact string matches were used to check for retrieval and hallucinations, hence codelists with formatting differences (e.g., missing the “^” at the start of codes) flagged in the assessment of failure modes may still be scored as correct in the main evaluation since the model-graded evaluation count formatting differences as mistakes.

| codelist | Epoch | Score | Retrieval |  |  |  | Hallucinations |  |  |
| --- | --- | --- | --- | --- | --- | --- | --- | --- | --- |
|  |  |  | Required | Retrieved | Not retrieved | Unused | Total | Hallucinated | % |
| Gemini 3 Pro |  |  |  |  |  |  |  |  |  |
| Atopic eczema | 1 | I | 56 | 54 | 2 | 0 | 55 | 0 | 0% |
|  | 2 | I | 56 | 47 | 9 | 0 | 49 | 1 | 2% |
|  | 3 | C | 56 | 56 | 0 | 0 | 58 | 0 | 0% |
| Vascular dementia | 1 | I | 63 | 36 | 27 | 0 | 72 | 29 | 40% |
|  | 2 | C | 63 | 37 | 26 | 0 | 67 | 0 | 0% |
|  | 3 | C | 63 | 37 | 26 | 0 | 72 | 0 | 0% |
| Eosinophilic esophagitis | 1 | C | 3 | 3 | 0 | 0 | 3 | 0 | 0% |
|  | 2 | C | 3 | 3 | 0 | 0 | 3 | 0 | 0% |
|  | 3 | C | 3 | 3 | 0 | 0 | 4 | 0 | 0% |
| Psoriasis | 1 | I | 76 | 74 | 2 | 0 | 85 | 0 | 0% |
|  | 2 | I | 76 | 74 | 2 | 0 | 87 | 0 | 0% |
|  | 3 | I | 76 | 74 | 2 | 0 | 91 | 0 | 0% |
| Hidradenitis suppurativa | 1 | C | 8 | 8 | 0 | 0 | 12 | 0 | 0% |
|  | 2 | C | 8 | 8 | 0 | 0 | 11 | 0 | 0% |
|  | 3 | C | 8 | 8 | 0 | 0 | 12 | 0 | 0% |
| Myocardial infarction | 1 | I | 132 | 70 | 62 | 0 | 93 | 1 | 1% |
|  | 2 | I | 132 | 113 | 19 | 113 | 0 | 0 | 0% |
|  | 3 | I | 132 | 102 | 30 | 102 | 0 | 0 | 0% |
| Wrist fracture | 1 | I | 214 | 74 | 140 | 74 | 0 | 0 | 0% |
|  | 2 | I | 214 | 104 | 110 | 3 | 113 | 1 | 1% |
|  | 3 | I | 214 | 123 | 91 | 0 | 136 | 2 | 1% |
| Claude Sonnet 4.6 |  |  |  |  |  |  |  |  |  |
| Atopic eczema | 1 | P | 56 | 56 | 0 | 0 | 97 | 8 | 8% |
|  | 2 | P | 56 | 56 | 0 | 0 | 63 | 3 | 5% |
|  | 3 | P | 56 | 56 | 0 | 0 | 80 | 7 | 9% |
| Vascular dementia | 1 | I | 64 | 37 | 27 | 1 | 74 | 8 | 11% |
|  | 2 | I | 64 | 37 | 27 | 0 | 73 | 3 | 4% |
|  | 3 | P | 64 | 38 | 26 | 0 | 72 | 5 | 7% |
| Eosinophilic esophagitis | 1 | C | 3 | 3 | 0 | 0 | 3 | 0 | 0% |
|  | 2 | C | 3 | 3 | 0 | 0 | 4 | 0 | 0% |
|  | 3 | C | 3 | 3 | 0 | 0 | 3 | 0 | 0% |
| Psoriasis | 1 | P | 76 | 74 | 2 | 0 | 89 | 7 | 8% |
|  | 2 | P | 76 | 76 | 0 | 1 | 112 | 12 | 11% |
|  | 3 | I | 76 | 73 | 3 | 1 | 85 | 6 | 7% |
| Hidradenitis suppurativa | 1 | P | 8 | 7 | 1 | 0 | 17 | 3 | 18% |
|  | 2 | C | 8 | 8 | 0 | 0 | 14 | 0 | 0% |
|  | 3 | I | 8 | 6 | 2 | 0 | 16 | 2 | 12% |
| Myocardial infarction | 1 | I | 132 | 95 | 37 | 95 | 0 | 0 | 0% |
|  | 2 | I | 132 | 108 | 24 | 108 | 0 | 0 | 0% |
|  | 3 | I | 132 | 122 | 10 | 122 | 0 | 0 | 0% |
| Wrist fracture | 1 | I | 214 | 89 | 125 | 89 | 0 | 0 | 0% |
|  | 2 | I | 214 | 95 | 119 | 95 | 0 | 0 | 0% |
|  | 3 | I | 214 | 131 | 83 | 131 | 0 | 0 | 0% |

We found that retrieval failures were present for all incorrect codelists across models. Generation failures (i.e., retrieved codelists not being used) were present in 4/12 (33%) for Gemini 3 Pro, 6/18 (33%) for GPT 5.2, 6/17 (35%) for Gemini 3 Flash, and 7/10 (70%) for Claude Sonnet 4.6. The list of not-retrieved and retrieved-but-not-used codes revealed variability across codelists and models (Supplementary Table 2).

### Alternative workflows

Using the string search workflow, we again saw varying accuracy across models with Gemini 3 Flash performing best on both the Full terminology (67%) and Read codes only (38%), followed by Claude Sonnet 4.6 (64% and 26%), and GPT 5.2 (55% and 14%). Failure patterns were different compared to the main evaluation. For example, misspelled codes included in the medical terminology such as “Hydradenitis suppurativa” were missed, as well as specific spelling variants like “radial” (not retrieved through a search for “radius”) or “infarct” (not retrieved through a search for “infarction”). (Supplementary Tables 4 & 5).

None of the other alternative workflows improved performance. Performance for the wrist fracture codelist did not improve when changing the number of [retrieved codes per tool call from 100 to 200](https://julianmatthewman.github.io/llmcodelists/#/logs/2026-01-30T11-39-57%2B00-00_RAG_f5178a12554e983adb7209.json/samples/sample/7/1/) (Score 0%; 3/3 incorrect; we only performed this evaluation with Gemini 3 Pro for the Read codes only evaluation for the wrist fracture codelist). Performance was worse when [using the MedCPT medical embedding model](https://julianmatthewman.github.io/llmcodelists/#/logs/2026-03-05T16-10-13%2B00-00_RAG_5ab6567c11a9be5c54356c.json) (30%) for Gemini 3 Pro (full terminology). (Correct: eosinophilic esophagitis and hidradenitis suppurativa in 3/3 epochs; partially correct: psoriasis in 1/3 epochs; incorrect: 14/21 all other epochs and codelists). An agentic workflow using Claude Code scored 21% (Correct: Eosinophilic esophagitis and psoriasis in 2/3 epochs; partially correct: atopic eczema in 1/3 epochs; incorrect: 16/21 all other epochs and codelists).

## Discussion and Conclusions

In summary, we found that retrieval augmented generation using current frontier large language models did create correct clinical codelists in some cases, but performance with the large and complex terminology was far from perfect, particularly with codelists containing a large number of codes.

A primary strength of this study is the development of bespoke reference codelists that capture the inherent ambiguity present when creating codelists with incomplete specifications. Creating bespoke reference codelists was also required since validated codelists for a given CPRD build are generally not publicly available or go out of date quickly. This is because the CPRD medical terminology consists of a mix of Read, SNOMED and local EMIS codes, with updates happening at each database release. We chose to evaluate the use of LLMs in this challenging scenario as it may be particularly useful as manually creating codelists for frequently changing and non-standardised terminologies often requires large ad-hoc effort.

Another strength of our study is that, beyond scoring the outputs, we detail specific failure modes, provide summaries, and test alternative methodological approaches. Our evaluation revealed varying accuracy across models but also highlighted model-specific failure profiles. Gemini 3 Pro and OpenAI GPT 5.2 generally had the lowest proportion of epochs containing hallucinations at 29%. In contrast, Claude Sonnet 4.6 exhibited the highest hallucination rate at 52%.

Required codes being retrieved at the retrieval step did not guarantee their inclusion in the final codelist produced in the generation step. We observed generation failures (where retrieved codes were not used by the model) in 70% of the evaluated incorrect epochs for Claude Sonnet, and approximately 33% for both Gemini 3 Pro and GPT 5.2. This indicates that improving the retrieval mechanism is insufficient if frontier models remain unable to fully synthesize and apply the retrieved context.

Since we performed the assessment of failure modes using exact string matching, hallucinations need to be distinguished from formatting discrepancies. We observed that some codes marked as being hallucinations were differences in formatting rather than wrong codes. For example, leaving out the “^” at the start of a local EMIS code resulted in a mismatch, though these were treated as correct codes during scoring. This highlights a practical challenge that LLMs may fail to output the exact string format required for direct database extraction. Future research should perform further evaluations on code-based approaches, as we did in our evaluation of Claude code, which generally avoid these types of hallucinations (but may suffer from other issues).

Our scoring was deliberately optimised for specificity, i.e., to minimise the chance of falsely scoring LLM outputs as incorrect. Firstly, we allowed for partially correct scores for codelists that capture all required codes even if they contained irrelevant codes. Secondly, we iterated on the reference codelists after seeing the evaluation results to correct any errors or omissions in the reference codelists. Despite the lenient scoring and iterated-upon reference codelists, performance was poor. We acknowledge that due to optimising specificity, scores may reflect an upper bound of LLM capabilities for the task.

We transparently report all evaluations which allows review of the evaluation process. We also highlight some of the unambiguous errors in our summaries and evaluation of failure modes, such as including foot fractures in the wrist fracture codelist, including codes that do not exist in the original code browser, or omitting codes that should unambiguously be included, like missing “heart attack” in the myocardial infarction codelist.

Even when required and optional codes are tagged, unambiguous reference standards may in general not be possible with simple specifications (i.e., just the disease name and the system prompt specifying that the codelists should capture current disease). For example, existing guidance on codelist development suggests a precise specification of the clinical concept, including timeframes, required accuracy of the codelist, setting, synonyms and exceptions.(1)

An example of ambiguity in our reference codelists is whether palmoplantar pustulosis should be included in the psoriasis codelists and whether a record would indicate the same conditions as a record for palmoplantar pustular psoriasis. There is no universal consensus in the dermatological profession whether these conditions should be considered the same entity.(15) In our evaluation “palmoplantar pustulosis” was not on the references codelist while “palmoplantar pustular psoriasis” was required on the reference codelist.

While we performed an evaluation on Read codes only, which considerably improved performance, further chunking may have improved performance even more. However, this may have also introduced problems since the mix of Read, SNOMED, and local EMIS codes in CPRD Aurum do not present a clear hierarchy in which to chunk codes.

An alternative to retrieval may be including the entire terminology in a model’s context window. While the context windows of some frontier models at the time of writing are large enough to fit all Read codes, there is degradation with longer contexts.(16) The maximum effective context window is generally much lower than the maximum context window, hence, RAG may remain superior to including the entire browser directly in the context window for now.

It must be acknowledged that in complex scenarios where correctness is essential, codelist development is an iterative process.(1) Iteration occurs both during the creation of a draft codelist as well as through expert review that helps clarify the codelist specification. Single-shot approaches, such as those tested here, may provide a useful starting point but may not be sufficient.

Despite the failures we demonstrate in this study, LLMs may be used for codelist development in a variety of ways. For example, LLMs and searches based on semantic similarity may serve as a basis to find relevant synonyms for a clinical concept, LLMs may assist in the review process, and, as we demonstrated, may produce correct codelists more frequently when the target terminology or codelist are smaller. We can anticipate ongoing performance improvement with AI, and we encourage future work that integrates the use of LLMs that can mitigate failures while harnessing the undeniable usefulness for the task of codelist development. Given the imperfect performance, guardrails would be required for routine trusted use.(17)

## Ethics

Ethical approval was not required for this study. This research did not involve human subjects and is exempt from Institutional Review Board or ethics committee oversight. The study methodology exclusively utilised a medical terminology and did not access, analyse, or generate any patient-level data.

## Author contributions

JM contributed to Conceptualization, Data curation, Formal analysis, Funding acquisition, Investigation, Methodology, Project administration, Resources, Software, Validation, Visualization, Writing – original draft, and Writing – review & editing.

AB contributed to Conceptualization, Methodology, Supervision, Validation, and Writing – review & editing.

SD, SML, JP contributed to Methodology, Supervision, Validation, and Writing – review & editing.

## List of Supplementary Tables

Supplementary Table 1: Hallucinations

Supplementary Table 2: Retrieved codes

Supplementary Table 3: Summary of failure modes for all models

Supplementary Table 4: AI-generated summaries of missed codes from string-search evaluation

Supplementary Table 5: AI-generated summaries of irrelevant codes from string-search evaluation

## References

1. Matthewman J, Andresen K, Suffel A, Lin LY, Schultze A, Tazare J, et al. Checklist and guidance on creating codelists for routinely collected health data research. NIHR Open Res. 2024 Sep 18;4:20. doi:10.3310/nihropenres.13550.2

2. Aslam A, Walker L, Abaho M, Cant H, O’Connell M, Abuzour AS, et al. An automation framework for clinical codelist development validated with UK data from patients with multiple long-term conditions. BMC Med Res Methodol. 2025 May 24;25(1):138. doi:10.1186/s12874-025-02541-1 PubMed PMID: 40413381; PubMed Central PMCID: PMC12102889.

3. Hoxhaj V, Riera-Arnau J, Mohammadi S, Sturkenboom MC, Navarro CLA. CodeMergeR: A shiny-based application for codelist integration [Internet]. medRxiv; 2025 [cited 2026 Apr 1]. p. 2025.12.07.25341784. Available from: https://www.medrxiv.org/content/10.64898/2025.12.07.25341784v1 doi:10.64898/2025.12.07.25341784

4. Becker BFH, Avillach P, Romio S, van Mulligen EM, Weibel D, Sturkenboom MCJM, et al. CodeMapper: semiautomatic coding of case definitions. A contribution from the ADVANCE project. Pharmacoepidemiol Drug Saf. 2017 Aug;26(8):998–1005. doi:10.1002/pds.4245 PubMed PMID: 28657162; PubMed Central PMCID: PMC5575526.

5. Singhal K, Azizi S, Tu T, Mahdavi SS, Wei J, Chung HW, et al. Large language models encode clinical knowledge. Nature. 2023 Aug;620(7972):172–80. doi:10.1038/s41586-023-06291-2

6. Yan C, Ong HH, Grabowska ME, Krantz MS, Su WC, Dickson AL, et al. Large language models facilitate the generation of electronic health record phenotyping algorithms. Journal of the American Medical Informatics Association. 2024 Sep 1;31(9):1994–2001. doi:10.1093/jamia/ocae072

7. Park H, Rees M, Kruger N, Fuse K, Castro VM, Gainer V, et al. Evaluation of T2DM Phenotyping Using Optimized Retrieval-Augmented Generation (RAG) and the Impact of Embedding Model, Context, and Prompt [Internet]. medRxiv; 2025 [cited 2026 Mar 10]. p. 2025.04.29.25326696. Available from: https://www.medrxiv.org/content/10.1101/2025.04.29.25326696v4 doi:10.1101/2025.04.29.25326696

8. Amugongo LM, Mascheroni P, Brooks S, Doering S, Seidel J. Retrieval augmented generation for large language models in healthcare: A systematic review. PLOS Digital Health. 2025 Jun 11;4(6):e0000877. doi:10.1371/journal.pdig.0000877

9. CPRD Aurum [Internet]. 2025 [cited 2026 Apr 1]. Available from: https://www.cprd.com/data/primary-care-data/cprd-aurum

10. England NHS. NHS England » Clinical coding – SNOMED CT [Internet]. [cited 2026 Apr 1]. Available from: https://www.england.nhs.uk/long-read/clinical-coding-snomed-ct/

11. Wickham H, Cheng J, Jacobs A, Aden-Buie G, Schloerke B. ellmer: Chat with Large Language Models [Internet]. 2025. Available from: https://ellmer.tidyverse.org

12. Kalinowski T, Falbel D. ragnar: Retrieval-Augmented Generation (RAG) Workflows [Internet]. 2026. Available from: https://ragnar.tidyverse.org/

13. Couch S. vitals: Large Language Model Evaluation [Internet]. 2025. Available from: https://github.com/tidyverse/vitals

14. Jin Q, Kim W, Chen Q, Comeau DC, Yeganova L, Wilbur WJ, et al. MedCPT: Contrastive Pre-trained Transformers with large-scale PubMed search logs for zero-shot biomedical information retrieval. Bioinformatics. 2023 Nov 1;39(11):btad651. doi:10.1093/bioinformatics/btad651

15. Yamamoto T. Similarity and difference between palmoplantar pustulosis and pustular psoriasis. The Journal of Dermatology. 2021;48(6):750–60. doi:10.1111/1346-8138.15826

16. Paulsen N. Context Is What You Need: The Maximum Effective Context Window for Real World Limits of LLMs [Internet]. arXiv; 2025 [cited 2026 Feb 6]. Available from: http://arxiv.org/abs/2509.21361 doi:10.48550/arXiv.2509.21361

17. Hakim JB, Painter JL, Ramcharran D, Kara V, Powell G, Sobczak P, et al. The need for guardrails with large language models in pharmacovigilance and other medical safety critical settings. Sci Rep. 2025 Jul 31;15(1):27886. doi:10.1038/s41598-025-09138-0

