## Supplementary Tables for "Large language models and retrieval augmented generation for complex clinical codelists: evaluating performance and assessing failure modes": Supplementary Table 1.html

| codelist | invalid\_codes |
| --- | --- |
| gemini-3-pro-preview | |
| --- | --- |
| Atopic eczema |  |
| Atopic eczema | ^ESCTDI512289 Discoid pattern atopic eczema |
| Atopic eczema |  |
| Vascular dementia | E004. Arteriosclerotic dementia, EMISICD10\|F0100 Vascular dementia of acute onset, without additional symptoms, EMISICD10\|F0101 Vascular dementia of acute onset, other symptoms, predominantly delusional, EMISICD10\|F0104 Vascular dementia of acute onset, other mixed symptoms, EMISICD10\|F0111 Multi-infarct dementia, other symptoms, predominantly delusional, EMISICD10\|F0113 Multi-infarct dementia, other symptoms, predominantly depressive, EMISICD10\|F0114 Multi-infarct dementia, other mixed symptoms, EMISICD10\|F0120 Subcortical vascular dementia, without additional symptoms, EMISICD10\|F0121 Subcortical vascular dementia, other symptoms, predominantly delusional, EMISICD10\|F0122 Subcortical vascular dementia, other symptoms, predominantly hallucinatory, EMISICD10\|F0123 Subcortical vascular dementia, other symptoms, predominantly depressive, EMISICD10\|F0124 Subcortical vascular dementia, other mixed symptoms, EMISICD10\|F0130 Mixed cortical and subcortical vascular dementia, without additional symptoms, EMISICD10\|F0131 Mixed cortical and subcortical vascular dementia, other symptoms, predominantly delusional, EMISICD10\|F0132 Mixed cortical and subcortical vascular dementia, other symptoms, predominantly hallucinatory, EMISICD10\|F0133 Mixed cortical and subcortical vascular dementia, other symptoms, predominantly depressive, EMISICD10\|F0134 Mixed cortical and subcortical vascular dementia, other mixed symptoms, EMISICD10\|F0180 Other vascular dementia, without additional symptoms, EMISICD10\|F0181 Other vascular dementia, other symptoms, predominantly delusional, EMISICD10\|F0182 Other vascular dementia, other symptoms, predominantly hallucinatory, EMISICD10\|F0183 Other vascular dementia, other symptoms, predominantly depressive, EMISICD10\|F0184 Other vascular dementia, other mixed symptoms, EMISICD10\|F0190 Vascular dementia, unspecified, without additional symptoms, EMISICD10\|F0191 Vascular dementia, unspecified, other symptoms, predominantly delusional, EMISICD10\|F0192 Vascular dementia, unspecified, other symptoms, predominantly hallucinatory, EMISICD10\|F0193 Vascular dementia, unspecified, other symptoms, predominantly depressive, EMISICD10\|F0194 Vascular dementia, unspecified, other mixed symptoms, Eu01. Vascular dementia, ^ESCT240788 Vascular dementia in remission |
| Vascular dementia |  |
| Vascular dementia |  |
| Eosinophilic esophagitis |  |
| Eosinophilic esophagitis |  |
| Eosinophilic esophagitis |  |
| Psoriasis |  |
| Psoriasis |  |
| Psoriasis | ^ESCTVУ666362 Vulval psoriasis |
| Hidradenitis suppurativa |  |
| Hidradenitis suppurativa |  |
| Hidradenitis suppurativa |  |
| Myocardial infarction | G27468 Subsequent myocardial infarction of anterior wall |
| Myocardial infarction |  |
| Myocardial infarction |  |
| Wrist fracture |  |
| Wrist fracture | ^ESCTMO435827 Monteggia's fracture (Often involves proximal ulna/radial head, but included if coded as wrist region in specific datasets, though usually elbow. S2341-99 is explicit Colles) |
| Wrist fracture | S4C03 Closed fracture-dislocation inferior radioulnar joint, ^ESCTCL484892 Closed fracture dislocation radioulnar joint |
| gpt-5.2 | |
| Atopic eczema |  |
| Atopic eczema |  |
| Atopic eczema |  |
| Vascular dementia |  |
| Vascular dementia |  |
| Vascular dementia | Eu011 [X] Multi-infarct dementia, Eu01y [X] Other vascular dementia, Eu01z [X] Vascular dementia, unspecified, Eu01-1 [X] Arteriosclerotic dementia |
| Eosinophilic esophagitis |  |
| Eosinophilic esophagitis |  |
| Eosinophilic esophagitis |  |
| Psoriasis | Myu30 [X] Other psoriasis, ^ESCT142366 Acute palmoplantar pustular psoriasis, ^ESCT149855 Chronic palmoplantar pustular psoriasis |
| Psoriasis |  |
| Psoriasis | ^ESCTPS666364 Penile psoriasis, ^ESCT157049 Psoriasiform dermatitis |
| Hidradenitis suppurativa |  |
| Hidradenitis suppurativa |  |
| Hidradenitis suppurativa |  |
| Myocardial infarction |  |
| Myocardial infarction |  |
| Myocardial infarction | G34 Subsequent myocardial infarction, Gyu35 [X] Subsequent myocardial infarction of other sites, Gyu36 [X] Subsequent myocardial infarction of unspecified site, Gyu34 [X] Acute transmural myocardial infarction of unspecif site |
| Wrist fracture |  |
| Wrist fracture |  |
| Wrist fracture | ^ESCTFR141933 Fracture of lunate bone of wrist |
| claude-sonnet-4-6 | |
| Atopic eczema | ^ESCTAT666132 Atopic eczema of face, ^ESCTAT666132 Atopic eczema of face, ^ESCTAT666132 Atopic eczema of face, ^ESCTAT666132 Atopic eczema of face, ^ESCTAT668133 Atopic dermatitis of eyelid, ^ESCTAC598664 Eczema of leg, Myu2 [X] Dermatitis and eczema, ^ESCT1246681 [X] Dermatitis and eczema |
| Atopic eczema | M111-99 Atopic eczema/dermatitis NOS, ^ESCTAT666131 Atopic dermatitis of eyelids, ^ESCTPS512296 Psoriasiform atopic eczema |
| Atopic eczema | M111-99 Atopic eczema/dermatitis NOS, ^ESCTAD666130 Atopic dermatitis of scalp (via ^ESCTAT666131 linked synonym), ^ESCTAD666132 Atopic dermatitis of face, ^ESCTAD666134 Atopic dermatitis of eyelid, ^ESCTAD666136 Atopic dermatitis of eyelids, ^ESCTAT666143 Adult atopic dermatitis, ^ESCTGE666370 Atopic dermatitis of eyelid |
| Vascular dementia | Eu011 [X] Multi-infarct dementia, Eu01y [X] Other vascular dementia, Eu01z [X] Vascular dementia, unspecified, Eu01-1 [X] Arteriosclerotic dementia, Eu011-1 [X] Predominantly cortical dementia, F5074 Binswanger's disease, ^ESCT1472649 Behavioural disturbance due to multi-infarct dementia, F26227 Cerebral degeneration due to cerebrovascular disease |
| Vascular dementia | F5074 Binswanger's disease (F21y2), ^ESCT1272649 Behavioural disturbance due to multi-infarct dementia, ^ESCTMU185163 Multi-infarct state |
| Vascular dementia | Eu011 [X] Multi-infarct dementia, Eu01y [X] Other vascular dementia, Eu01z [X] Vascular dementia, unspecified, Eu01-1 [X] Arteriosclerotic dementia, Eu011-1 [X] Predominantly cortical dementia |
| Eosinophilic esophagitis |  |
| Eosinophilic esophagitis |  |
| Eosinophilic esophagitis |  |
| Psoriasis | Myu30 [X] Other psoriasis, ^ESCTVÜ666362 Vulval psoriasis, ^ESCTPS512395 Psoriatic nail pitting, ^ESCTPS512394 Psoriatic onycholysis, ^ESCTGE311787 Generalised pustular psoriasis of von Zumbusch, ^ESCTJU512406 Infantile pustular psoriasis, ^ESCTAC666403 Acrodermatitis continua of Hallopeau |
| Psoriasis | ^ESCTGE666345 Generalised psoriasis, ^ESCTPS512395 Psoriatic nail pitting, ^ESCTSE291360 Seborrhoeic psoriasis, ^ESCTPS512394 Psoriatic onycholysis, ^ESCTAC166236 Acrodermatitis continua of Hallopeau, ^ESCTIR450796 Iritis in psoriatic arthritis, ^ESCTIR230450 Iritis in psoriatic arthritis, ^ESCTOL453163 Oligoarticular psoriatic arthritis, ^ESCTMU513961 Psoriatic arthritis mutilans |
| Psoriasis | Myu30 [X] Other psoriasis, ^ESCTPS512395 Psoriatic nail pitting, ^ESCTPS512394 Psoriatic onycholysis (nail), ^ESCTJU512406 Infantile pustular psoriasis, ^ESCTCH666351 Palmoplantar pustulosis, M143 Impetigo herpetiformis (pustular psoriasis of pregnancy) |
| Hidradenitis suppurativa | ^ESCTVУ667049 Vulval hidradenitis suppurativa, ^ESCTFO669402 / ^ESCTFO669403 Follicular occlusion triad - hidradenitis, acne conglobata, dissecting cellulitis of scalp, Myu6D [X] Other apocrine sweat disorders |
| Hidradenitis suppurativa |  |
| Hidradenitis suppurativa | ^ESCTVУ667049 Vulval hidradenitis suppurativa, Myu6D [X] Other apocrine sweat disorders |
| Myocardial infarction |  |
| Myocardial infarction |  |
| Myocardial infarction |  |
| Wrist fracture |  |
| Wrist fracture |  |
| Wrist fracture |  |
| gemini-3-flash-preview | |
| Atopic eczema |  |
| Atopic eczema | ESCTAT288430 Atopic eczema, ESCTAD288434 AD - Atopic dermatitis, ESCTCH666142 Childhood atopic eczema, ESCTIN666140 Infantile atopic eczema, ESCTAD666144 Adult atopic eczema, ESCTGE666122 Generalised atopic eczema, ESCTAT666133 Atopic eczema of face, ESCTER512292 Erythrodermic atopic eczema, ESCTCH666141 Childhood atopic dermatitis, ESCTAT666131 Atopic eczema of scalp, ESCTAL288431 Allergic eczema, ESCTPH512299 Photosensitive atopic eczema, ESCTPH512301 Photoaggravated atopic eczema, ESCTCH598918 Chronic lichenified atopic eczema, ESCTFL666117 Flexural atopic eczema, ESCTAD666143 Adult atopic dermatitis, ESCTPR512296 Pruriginous atopic eczema, ESCTGE666120 Generalised atopic dermatitis, ESCTIN666139 Infantile atopic dermatitis, ESCTAD666146 Adult atopic eczema persistent from childhood, ESCT1500750 Childhood generalised erythematous atopic dermatitis, ESCTER512291 Erythrodermic atopic dermatitis, ESCTCH666138 Cheilitis due to atopic eczema, ESCTAT512286 Atopic eczema of hands, ESCTAT666134 Atopic dermatitis of eyelid, ESCTDI512290 Discoid pattern atopic eczema, ESCTAT666132 Atopic dermatitis of face, ESCTAD666148 Adult atopic eczema recurrent in adult life, ESCTAT666136 Atopic dermatitis of eyelids, ESCTCH598917 Chronic lichenified atopic dermatitis, ESCTAC267510 Acute infantile eczema, ESCTFO512294 Follicular atopic eczema, ESCTPH512297 Photosensitive atopic dermatitis, ESCTAT666130 Atopic dermatitis of scalp, ESCTFL666116 Flexural atopic dermatitis, ESCTIN666152 Infected atopic dermatitis, ESCTPH512300 Photoaggravated atopic dermatitis, ESCT1450246 Atopic dermatitis of bilateral hands, ESCTIM666151 Impetiginized atopic dermatitis, ESCT1450704 Acute atopic dermatitis of hand, ESCTAT512285 Atopic dermatitis of hands, ESCTPR666124 Prurigo pattern atopic dermatitis, ESCTDI512288 Discoid atopic dermatitis, ESCTDI512289 Discoid pattern atopic dermatitis, ESCTIN512287 Inverse pattern atopic dermatitis |
| Atopic eczema |  |
| Vascular dementia |  |
| Vascular dementia | EMISICD10\|F0180 Other vascular dementia, without additional symptoms, EMISICD10\|F0181 Other vascular dementia, other symptoms, predominantly delusional, EMISICD10\|F0191 Vascular dementia, unspecified, other symptoms, predominantly delusional, EMISICD10\|F0100 Vascular dementia of acute onset, without additional symptoms, EMISICD10\|F0104 Vascular dementia of acute onset, other mixed symptoms, EMISICD10\|F0184 Other vascular dementia, other mixed symptoms, EMISICD10\|F0183 Other vascular dementia, other symptoms, predominantly depressive, EMISICD10\|F0190 Vascular dementia, unspecified, without additional symptoms, EMISICD10\|F0193 Vascular dementia, unspecified, other symptoms, predominantly depressive, EMISICD10\|F0101 Vascular dementia of acute onset, other symptoms, predominantly delusional, EMISICD10\|F0194 Vascular dementia, unspecified, other mixed symptoms, EMISICD10\|F0124 Subcortical vascular dementia, other mixed symptoms, EMISICD10\|F0120 Subcortical vascular dementia, without additional symptoms, EMISICD10\|F0121 Subcortical vascular dementia, other symptoms, predominantly delusional, EMISICD10\|F0123 Subcortical vascular dementia, other symptoms, predominantly depressive, EMISICD10\|F0182 Other vascular dementia, other symptoms, predominantly hallucinatory, EMISICD10\|F0130 Mixed cortical and subcortical vascular dementia, without additional symptoms, EMISICD10\|F0192 Vascular dementia, unspecified, other symptoms, predominantly hallucinatory, EMISICD10\|F0134 Mixed cortical and subcortical vascular dementia, other mixed symptoms, EMISICD10\|F0131 Mixed cortical and subcortical vascular dementia, other symptoms, predominantly delusional, EMISICD10\|F0133 Mixed cortical and subcortical vascular dementia, other symptoms, predominantly depressive, EMISICD10\|F0122 Subcortical vascular dementia, other symptoms, predominantly hallucinatory, EMISICD10\|F0132 Mixed cortical and subcortical vascular dementia, other symptoms, predominantly hallucinatory, EMISICD10\|F0114 Multi-infarct dementia, other mixed symptoms, EMISICD10\|F0111 Multi-infarct dementia, other symptoms, predominantly delusional, EMISICD10\|F0113 Multi-infarct dementia, other symptoms, predominantly depressive |
| Vascular dementia | 341425000 VAD - Vascular dementia, 291233008 Vascular dementia, with delusions, 266461001 Vascular dementia, with delirium, 990225008 Vascular dementia without behavioural disturbance, 808924009 Vascular dementia with behavioral disturbance, 808923003 Vascular dementia with behavioural disturbance, 784063001 Ischaemic vascular dementia, 784064007 Ischemic vascular dementia, 1393438006 Cortical vascular dementia, 1171774004 Predominantly cortical vascular dementia, 1503911000 Dementia due to cerebral vasculitis, 341423007 MID - Multi-infarct dementia, 341426006 Multi infarct dementia, 365367008 Multi-infarct dementia, uncomplicated, 266460000 Multi-infarct dementia with delirium, 803324005 Multi-infarct dementia due to atherosclerosis, 396466004 Subcortical atherosclerotic dementia, 1237065005 MVAD - Mixed vascular Alzheimer dementia, F0100 Vascular dementia of acute onset, without additional symptoms, F0101 Vascular dementia of acute onset, other symptoms, predominantly delusional, F0104 Vascular dementia of acute onset, other mixed symptoms, F0111 Multi-infarct dementia, other symptoms, predominantly delusional, F0113 Multi-infarct dementia, other symptoms, predominantly depressive, F0114 Multi-infarct dementia, other mixed symptoms, F0120 Subcortical vascular dementia, without additional symptoms, F0121 Subcortical vascular dementia, other symptoms, predominantly delusional, F0122 Subcortical vascular dementia, other symptoms, predominantly hallucinatory, F0123 Subcortical vascular dementia, other symptoms, predominantly depressive, F0124 Subcortical vascular dementia, other mixed symptoms, F0130 Mixed cortical and subcortical vascular dementia, without additional symptoms, F0131 Mixed cortical and subcortical vascular dementia, other symptoms, predominantly delusional, F0132 Mixed cortical and subcortical vascular dementia, other symptoms, predominantly hallucinatory, F0133 Mixed cortical and subcortical vascular dementia, other symptoms, predominantly depressive, F0134 Mixed cortical and subcortical vascular dementia, other mixed symptoms, F0180 Other vascular dementia, without additional symptoms, F0181 Other vascular dementia, other symptoms, predominantly delusional, F0182 Other vascular dementia, other symptoms, predominantly hallucinatory, F0183 Other vascular dementia, other symptoms, predominantly depressive, F0184 Other vascular dementia, other mixed symptoms, F0190 Vascular dementia, unspecified, without additional symptoms, F0191 Vascular dementia, unspecified, other symptoms, predominantly delusional, F0192 Vascular dementia, unspecified, other symptoms, predominantly hallucinatory, F0193 Vascular dementia, unspecified, other symptoms, predominantly depressive, F0194 Vascular dementia, unspecified, other mixed symptoms |
| Eosinophilic esophagitis |  |
| Eosinophilic esophagitis | ESCTEO508265 Eosinophilic esophagitis, ESCT1165474 Food-induced eosinophilic oesophagitis, ESCTEO508286 Eosinophilic ulcer of oesophagus |
| Eosinophilic esophagitis |  |
| Psoriasis | ^ESCTFL1 Flexural psoriasis |
| Psoriasis |  |
| Psoriasis |  |
| Hidradenitis suppurativa |  |
| Hidradenitis suppurativa |  |
| Hidradenitis suppurativa |  |
| Myocardial infarction | ^ESCT323 ECG: myocardial infarction |
| Myocardial infarction |  |
| Myocardial infarction | G3071 Acute non-ST segment elevation myocardial infarction (NSTEMI), G30X Acute transmural myocardial infarction of unspecified site, G30X0 Acute ST segment elevation myocardial infarction (STEMI) |
| Wrist fracture | S234B Closed fracture radial styloid (Chauffeur's) |
| Wrist fracture |  |
| Wrist fracture | S234B Closed fracture radial styloid (Chauffeur's fracture), ^ESCTCL435380 Closed reverse Colles' fracture (Smith's) |
