## Supplementary Tables for "Large language models and retrieval augmented generation for complex clinical codelists: evaluating performance and assessing failure modes": Supplementary Table 2.html

| Epoch | Score | Not retrieved | Retrieved but unused |
| --- | --- | --- | --- |
| Atopic eczema - gemini-3-pro-preview | | | |
| --- | --- | --- | --- |
| 1 | I | M115, ^ESCTXE666127 |  |
| 2 | I | ^ESCTDI512288, ^ESCTDI512290, ^ESCTFO512293, ^ESCTPR512295, ^ESCTPH512297, ^ESCTPH512299, ^ESCTXE666127, ^ESCTCH666138, ^ESCTAD666143 |  |
| Vascular dementia - gemini-3-pro-preview | | | |
| 1 | I | E004-1, EMISICD10\|F0100, EMISICD10\|F0101, EMISICD10\|F0104, EMISICD10\|F0111, EMISICD10\|F0113, EMISICD10\|F0114, EMISICD10\|F0120, EMISICD10\|F0121, EMISICD10\|F0122, EMISICD10\|F0123, EMISICD10\|F0124, EMISICD10\|F0130, EMISICD10\|F0131, EMISICD10\|F0132, EMISICD10\|F0133, EMISICD10\|F0134, EMISICD10\|F0180, EMISICD10\|F0181, EMISICD10\|F0182, EMISICD10\|F0183, EMISICD10\|F0184, EMISICD10\|F0190, EMISICD10\|F0191, EMISICD10\|F0192, EMISICD10\|F0193, EMISICD10\|F0194 |  |
| 2 | C | EMISICD10\|F0100, EMISICD10\|F0101, EMISICD10\|F0104, EMISICD10\|F0111, EMISICD10\|F0113, EMISICD10\|F0114, EMISICD10\|F0120, EMISICD10\|F0121, EMISICD10\|F0122, EMISICD10\|F0123, EMISICD10\|F0124, EMISICD10\|F0130, EMISICD10\|F0131, EMISICD10\|F0132, EMISICD10\|F0133, EMISICD10\|F0134, EMISICD10\|F0180, EMISICD10\|F0181, EMISICD10\|F0182, EMISICD10\|F0183, EMISICD10\|F0184, EMISICD10\|F0190, EMISICD10\|F0191, EMISICD10\|F0192, EMISICD10\|F0193, EMISICD10\|F0194 |  |
| 3 | C | EMISICD10\|F0100, EMISICD10\|F0101, EMISICD10\|F0104, EMISICD10\|F0111, EMISICD10\|F0113, EMISICD10\|F0114, EMISICD10\|F0120, EMISICD10\|F0121, EMISICD10\|F0122, EMISICD10\|F0123, EMISICD10\|F0124, EMISICD10\|F0130, EMISICD10\|F0131, EMISICD10\|F0132, EMISICD10\|F0133, EMISICD10\|F0134, EMISICD10\|F0180, EMISICD10\|F0181, EMISICD10\|F0182, EMISICD10\|F0183, EMISICD10\|F0184, EMISICD10\|F0190, EMISICD10\|F0191, EMISICD10\|F0192, EMISICD10\|F0193, EMISICD10\|F0194 |  |
| Psoriasis - gemini-3-pro-preview | | | |
| 1 | I | ^ESCTPS512393, ^ESCTPS512395 |  |
| 2 | I | ^ESCTPS512393, ^ESCTPS512395 |  |
| 3 | I | ^ESCTPS512393, ^ESCTPS512395 |  |
| Myocardial infarction - gemini-3-pro-preview | | | |
| 1 | I | G3010, G30z, G303, G30X0, G3071, Gyu34, ^ESCTAC345218, ^ESCTAC364164, ^ESCTAC378491, ^ESCTAC505634, ^ESCTNO604377, ^ESCTNS665139, ^ESCTAC757158, ^ESCTAC757161, ^ESCTAC757162, ^ESCTAC757232, ^ESCTAC757234, ^ESCTAC757288, ^ESCTAC757290, ^ESCTAC808775, ^ESCTAC984395, ^ESCTAC984396, ^ESCTAC984399, ^ESCTAC984401, ^ESCTAC984405, ^ESCTAC984407, ^ESCT1212233, ^ESCT1212234, ^ESCT1393516, ^ESCT1393521, ^ESCT1394109, ^ESCT1394111, ^ESCT1394114, ^ESCT1394116, ^ESCT1394121, ^ESCT1394123, ^ESCT1394521, ^ESCT1394523, ^ESCT1394772, ^ESCT1396130, ^ESCT1396131, ^ESCT1396134, ^ESCT1396135, ^ESCT1396138, ^ESCT1396142, ^ESCT1396143, ^ESCT1396144, ^ESCT1396146, ^ESCT1396147, ^ESCT1413294, ^ESCT1413295, ^ESCT1414539, ^ESCT1414543, ^ESCT1414546, ^ESCT1474197, ^ESCT1474201, ^ESCT1474299, ^ESCT1527612, ^ESCT1527615, ^ESCT1527618, ^ESCT1527621, ^ESCT1527624 |  |
| 2 | I | ^ESCTNS665139, ^ESCTAC757158, ^ESCTAC757162, ^ESCTAC808775, ^ESCTAC984396, ^ESCTAC984402, ^ESCTAC984405, ^ESCT1212233, ^ESCT1212234, ^ESCT1393516, ^ESCT1394772, ^ESCT1396130, ^ESCT1396131, ^ESCT1396134, ^ESCT1396135, ^ESCT1396142, ^ESCT1413294, ^ESCT1414539, ^ESCT1527615 |  |
| 3 | I | G3010, G303, Gyu34, G302, ^ESCTAC334347, ^ESCTAC378491, ^ESCTAC757158, ^ESCTAC757234, ^ESCTAC801104, ^ESCTAC808775, ^ESCT1212234, ^ESCT1393516, ^ESCT1393519, ^ESCT1394109, ^ESCT1394114, ^ESCT1394121, ^ESCT1394521, ^ESCT1394772, ^ESCT1396130, ^ESCT1396134, ^ESCT1396142, ^ESCT1396144, ^ESCT1396146, ^ESCT1414537, ^ESCT1414543, ^ESCT1414546, ^ESCT1527612, ^ESCT1527618, ^ESCT1527621, ^ESCT1527624 |  |
| Wrist fracture - gemini-3-pro-preview | | | |
| 1 | I | S2341-1, S2351-1, S2359, S235A, S240z, S2410, S2419, S241F, S241y, S241z, S235F, S234F, S2406, S2416, S24-1, ^ESCT1167569, ^ESCT1167570, S2359-1, S2359-2, S4C31, S4C1y, S4C15, S4C16, S4C11, S4C3y, S4C14, S4C12, S235A-1, S235A-2, S234A, S234A-2, S234A-1, S4C0y, S4C22, S4C2y, S2349-2, S2349-1, S2341-98, S2341-99, ^ESCTCL250422, ^ESCTFR253074, ^ESCTFR253076, ^ESCTFR262068, ^ESCTFR262070, ^ESCTFR262071, ^ESCTOP262611, ^ESCTCL265096, ^ESCTFR273648, ^ESCTFR273650, ^ESCTCL277320, ^ESCTCL299359, ^ESCTFR301489, ^ESCTFR301491, ^ESCTCL303424, ^ESCTOP305807, ^ESCTCL309869, ^ESCTCL310603, ^ESCTCL319032, ^ESCTCL320474, ^ESCTOP321700, ^ESCTOP336361, ^ESCTBA338678, ^ESCTBA338679, ^ESCTCL342815, ^ESCTOP359139, ^ESCTOP360729, ^ESCTFR369224, ^ESCTFR369226, ^ESCTFR371033, ^ESCTFR371035, ^ESCTFR383451, ^ESCTFR386705, ^ESCTFR386706, ^ESCTFR389649, ^ESCTFR389651, ^ESCTOP393200, ^ESCTOP394930, ^ESCTCL396486, ^ESCTAR421400, ^ESCTCL435380, ^ESCTOP435381, ^ESCTCO435824, ^ESCTRE435825, ^ESCTSM435826, ^ESCTCL484225, ^ESCTCL484228, ^ESCTCL484229, ^ESCTCL484892, ^ESCTCL484893, ^ESCTCL484896, ^ESCTCL484897, ^ESCTCL484898, ^ESCTOP484902, ^ESCTOP484905, ^ESCTOP484906, ^ESCTOP484907, ^ESCTCL484910, ^ESCTCL484913, ^ESCTCL484914, ^ESCTCL484915, ^ESCTOP484916, ^ESCTOP484921, ^ESCTFR546059, ^ESCTFR546060, ^ESCTFR546062, ^ESCTFR546082, ^ESCTFR546088, ^ESCTFR546199, ^ESCTVO546201, ^ESCTVO546202, ^ESCTDO546203, ^ESCTDO546204, ^ESCTFR546212, ^ESCTTR565252, ^ESCTTR565254, ^ESCTTR565255, ^ESCTTR565256, ^ESCTFR565268, ^ESCTCL597198, ^ESCTPA730975, ^ESCTCL758786, ^ESCTPA766315, ^ESCT1221720, ^ESCT1370300, ^ESCT1370301, ^ESCT1370303, ^ESCT1379051, ^ESCT1419672, ^ESCT1507936, ^ESCT1507937, ^ESCT1526232, ^ESCT1529546, ^ESCT1543734, ^ESCT1543735, ^ESCT1543738, ^ESCT1565378, ^ESCT1605454, ^ESCT1608054, ^ESCTHU705948, ^ESCTCH705949 | S4C3, S4C |
| 2 | I | S2349, S2359, S235A, S235E, S2400, S240z, S2410, S2419, S241F, S241y, S241z, Syu63, S23B, S23C, S235F, S234F, S2416, S2356, ^ESCT1167569, ^ESCT1167570, S2359-1, S2359-2, S4C36, S4C31, S4C1y, S4C15, S4C16, S4C11, S4C3, S4C3y, S4C14, S4C12, S235A-1, S235A-2, Syu65, S234A, S234A-2, S234A-1, S4C04, S4C02, S4C0y, S4C05, S4C01, S4C24, S4C22, S4C2y, S4C25, S4C21, S2349-2, S2349-1, S234G, ^ESCTFR253076, ^ESCTFR262070, ^ESCTFR262071, ^ESCTOP262611, ^ESCTFR273650, ^ESCTFR301489, ^ESCTOP305807, ^ESCTOP321700, ^ESCTOP336361, ^ESCTBA338678, ^ESCTBA338679, ^ESCTOP359139, ^ESCTOP360729, ^ESCTFR369226, ^ESCTFR371035, ^ESCTFR386706, ^ESCTFR389651, ^ESCTOP393200, ^ESCTOP394930, ^ESCTAR421400, ^ESCTCL435380, ^ESCTOP435381, ^ESCTRE435825, ^ESCTCL484225, ^ESCTCL484228, ^ESCTCL484229, ^ESCTCL484892, ^ESCTCL484893, ^ESCTCL484896, ^ESCTCL484897, ^ESCTCL484898, ^ESCTOP484902, ^ESCTOP484905, ^ESCTOP484906, ^ESCTOP484907, ^ESCTCL484910, ^ESCTCL484913, ^ESCTCL484914, ^ESCTCL484915, ^ESCTOP484916, ^ESCTOP484921, ^ESCTFR546060, ^ESCTFR546062, ^ESCTFR546088, ^ESCTDO546203, ^ESCTDO546204, ^ESCTTR565254, ^ESCTTR565255, ^ESCTCL597198, ^ESCTPA730975, ^ESCTCL758786, ^ESCTPA766315, ^ESCT1221720, ^ESCT1370300, ^ESCT1370301, ^ESCT1370303, ^ESCT1419672, ^ESCT1507937, ^ESCTHU705948 | S242, S234, S235 |
| 3 | I | S2341-1, S234B, S2351-1, S240y, S2410, S241y, S241z, S24z, S4C1, S2406, S2416, S2411, ^ESCT1167569, ^ESCT1167570, S2359-1, S2359-2, S4C36, S4C31, S4C1y, S4C15, S4C16, S4C11, S4C3, S4C3y, S4C14, S4C12, S235A-1, S235A-2, S234A-2, S234A-1, S4C0y, S4C22, S4C2y, S4C21, S2349-2, S2349-1, S2341-98, S234G, ^ESCTFR253076, ^ESCTFR262070, ^ESCTFR262071, ^ESCTOP262611, ^ESCTCL265096, ^ESCTFR273650, ^ESCTOP305807, ^ESCTOP321700, ^ESCTOP336361, ^ESCTOP359139, ^ESCTOP360729, ^ESCTFR369226, ^ESCTFR371035, ^ESCTFR386706, ^ESCTFR389651, ^ESCTOP394930, ^ESCTAR421400, ^ESCTCL435380, ^ESCTOP435381, ^ESCTRE435825, ^ESCTCL484225, ^ESCTCL484228, ^ESCTCL484229, ^ESCTCL484896, ^ESCTCL484897, ^ESCTOP484902, ^ESCTOP484905, ^ESCTOP484906, ^ESCTOP484907, ^ESCTCL484910, ^ESCTCL484913, ^ESCTCL484914, ^ESCTCL484915, ^ESCTOP484916, ^ESCTOP484921, ^ESCTFR546060, ^ESCTFR546062, ^ESCTFR546088, ^ESCTVO546202, ^ESCTDO546204, ^ESCTFR546212, ^ESCTTR565255, ^ESCTCL597198, ^ESCTPA730975, ^ESCTCL758786, ^ESCTPA766315, ^ESCT1221720, ^ESCT1379051, ^ESCT1419672, ^ESCT1507937, ^ESCT1565378, ^ESCTHU705948, ^ESCTCH705949 |  |
| Atopic eczema - gpt-5.2 | | | |
| 1 | I | M113, M115, M114, ^ESCTIN512287, ^ESCTFO512293, ^ESCTPR512295, ^ESCTPR666124, ^ESCTIM666151 |  |
| 2 | I | M115, ^ESCTCH598917, ^ESCTCH598918, ^ESCT1500750 |  |
| 3 | I | M113, M115, M114, ^ESCTIN512287, ^ESCTFO512293, ^ESCTPR512295, ^ESCTPR666124, ^ESCTIM666151 |  |
| Vascular dementia - gpt-5.2 | | | |
| 1 | I | F21y2, E0041, E0042, E0043, F21y2-1, EMISICD10\|F0100, EMISICD10\|F0101, EMISICD10\|F0104, EMISICD10\|F0111, EMISICD10\|F0113, EMISICD10\|F0114, EMISICD10\|F0120, EMISICD10\|F0121, EMISICD10\|F0122, EMISICD10\|F0123, EMISICD10\|F0124, EMISICD10\|F0130, EMISICD10\|F0131, EMISICD10\|F0132, EMISICD10\|F0133, EMISICD10\|F0134, EMISICD10\|F0180, EMISICD10\|F0181, EMISICD10\|F0182, EMISICD10\|F0183, EMISICD10\|F0184, EMISICD10\|F0190, EMISICD10\|F0191, EMISICD10\|F0192, EMISICD10\|F0193, EMISICD10\|F0194, ^ESCTMU266460, ^ESCTMU272293, ^ESCTMU291232, ^ESCTSU396466, ^ESCTMU803324, ^ESCT1480525, ^ESCTSU396465 |  |
| 2 | I | F21y2, E0041, E0042, E0043, F21y2-1, EMISICD10\|F0100, EMISICD10\|F0101, EMISICD10\|F0104, EMISICD10\|F0111, EMISICD10\|F0113, EMISICD10\|F0114, EMISICD10\|F0120, EMISICD10\|F0121, EMISICD10\|F0122, EMISICD10\|F0123, EMISICD10\|F0124, EMISICD10\|F0130, EMISICD10\|F0131, EMISICD10\|F0132, EMISICD10\|F0133, EMISICD10\|F0134, EMISICD10\|F0180, EMISICD10\|F0181, EMISICD10\|F0182, EMISICD10\|F0183, EMISICD10\|F0184, EMISICD10\|F0190, EMISICD10\|F0191, EMISICD10\|F0192, EMISICD10\|F0193, EMISICD10\|F0194, ^ESCTMU266460, ^ESCTMU272293, ^ESCTMU291232, ^ESCTSU396466, ^ESCTMU803324, ^ESCT1480525, ^ESCTSU396465 |  |
| 3 | I | F21y2, E0041, E0042, E0043, F21y2-1, EMISICD10\|F0100, EMISICD10\|F0101, EMISICD10\|F0104, EMISICD10\|F0111, EMISICD10\|F0113, EMISICD10\|F0114, EMISICD10\|F0120, EMISICD10\|F0121, EMISICD10\|F0122, EMISICD10\|F0123, EMISICD10\|F0124, EMISICD10\|F0130, EMISICD10\|F0131, EMISICD10\|F0132, EMISICD10\|F0133, EMISICD10\|F0134, EMISICD10\|F0180, EMISICD10\|F0181, EMISICD10\|F0182, EMISICD10\|F0183, EMISICD10\|F0184, EMISICD10\|F0190, EMISICD10\|F0191, EMISICD10\|F0192, EMISICD10\|F0193, EMISICD10\|F0194, ^ESCTMU266460, ^ESCTMU272293, ^ESCTMU291232, ^ESCTSU396466, ^ESCTVA751506, ^ESCTMU803324, ^ESCT1480525, ^ESCTSU396465 | E0040, E004z, Eu01y, Eu01z, E004, Eu01-1, Eu011, Eu012, Eu01, Eu010, E004-1, EMISNQDV1, ^ESCTVA266461, ^ESCT1171774, ^ESCTVA291233, ^ESCTMI341423, ^ESCTVA341425, ^ESCTMU341426, ^ESCTMU365367, ^ESCTIS784063, ^ESCTIS784064, ^ESCTVA808923, ^ESCTVA808924, ^ESCTVA990225, ^ESCT1393438 |
| Psoriasis - gpt-5.2 | | | |
| 1 | I | M1612, M1613, M161C, M166, ^ESCTAC255528, ^ESCTCH294145, ^ESCTPS309677, ^ESCTNU480582, ^ESCTIN512386, ^ESCTPS512392, ^ESCTPS512393, ^ESCTPS512394, ^ESCTPS512395, ^ESCTPE666366, ^ESCTGE666367 | Nyu13 |
| 2 | I | M1612, Myu30, ESCTFL1, M166, M161J, ^ESCTAC255528, ^ESCTCH294145, ^ESCTPU294148, ^ESCTPS309677, ^ESCTGE311787, ^ESCTLO382192, ^ESCTNU480582, ^ESCTIN512386, ^ESCTUN512387, ^ESCTPS512388, ^ESCTEC512389, ^ESCTEC512390, ^ESCTPS512392, ^ESCTPS512393, ^ESCTPS512394, ^ESCTPS512395, ^ESCTKÖ512397, ^ESCTKO512398, ^ESCTGE512400, ^ESCTGE512402, ^ESCTPU512404, ^ESCTJU512405, ^ESCTIN512406, ^ESCTGU666360, ^ESCTPS666365, ^ESCTPE666366, ^ESCTHY666374, ^ESCTHY666375, ^ESCTAC666380, ^ESCTCH666382, ^ESCT1363707, ^ESCT1419756, ^ESCT1419757 |  |
| 3 | I | M161H, Myu30, ESCTFL1, M166, EMISNQCH31, M161F-1, M161J, ^ESCTAC255528, ^ESCTCH294145, ^ESCTPU294148, ^ESCTLO382192, ^ESCTIN512386, ^ESCTPS512393, ^ESCTPS512395, ^ESCTKO512398, ^ESCTGE512400, ^ESCTGE512402, ^ESCTPU512404, ^ESCTJU512405, ^ESCTIN512406, ^ESCTPE666364, ^ESCTPS666365, ^ESCTPE666366, ^ESCTHY666375, ^ESCTCH666382, ^ESCT1419758 |  |
| Hidradenitis suppurativa - gpt-5.2 | | | |
| 1 | I | ^ESCTVE346453 |  |
| 2 | I | ^ESCTVE346453 |  |
| 3 | I | ^ESCTVE346453 |  |
| Myocardial infarction - gpt-5.2 | | | |
| 1 | I | G30-4, G307, G3010, G304, G305, G306, G308, G30y0, G3070, G30y1, G303, G30y2, G30X0, G3071, G300, G3011, G302, G30X, G30-99, G30B, ^ESCTAC250300, ^ESCT1171766, ^ESCT1171768, ^ESCTAC334347, ^ESCTAC338160, ^ESCTAC345218, ^ESCTAC345956, ^ESCTAC355117, ^ESCTAC356587, ^ESCTAC364164, ^ESCTAC369992, ^ESCTAC374574, ^ESCTAC378491, ^ESCTAC477589, ^ESCTAC505627, ^ESCTAC505629, ^ESCTAC505630, ^ESCTAC505631, ^ESCTAC505632, ^ESCTAC505633, ^ESCTAC505634, ^ESCTAC593532, ^ESCTNO604377, ^ESCTST665122, ^ESCTNS665139, ^ESCTAC757158, ^ESCTAC757159, ^ESCTAC757160, ^ESCTAC757161, ^ESCTAC757162, ^ESCTAC757232, ^ESCTAC757233, ^ESCTAC757234, ^ESCTAC757286, ^ESCTAC757288, ^ESCTAC757290, ^ESCTAC800426, ^ESCTAC801104, ^ESCTAC808775, ^ESCTAC984395, ^ESCTAC984396, ^ESCTAC984398, ^ESCTAC984399, ^ESCTAC984401, ^ESCTAC984402, ^ESCTAC984405, ^ESCTAC984407, ^ESCT1212233, ^ESCT1212234, ^ESCT1393516, ^ESCT1393519, ^ESCT1393520, ^ESCT1393521, ^ESCT1394109, ^ESCT1394111, ^ESCT1394114, ^ESCT1394116, ^ESCT1394121, ^ESCT1394123, ^ESCT1394521, ^ESCT1394523, ^ESCT1394598, ^ESCT1394759, ^ESCT1394772, ^ESCT1396130, ^ESCT1396131, ^ESCT1396134, ^ESCT1396135, ^ESCT1396137, ^ESCT1396138, ^ESCT1396142, ^ESCT1396143, ^ESCT1396144, ^ESCT1396145, ^ESCT1396146, ^ESCT1396147, ^ESCT1401503, ^ESCT1413294, ^ESCT1413295, ^ESCT1414537, ^ESCT1414539, ^ESCT1414543, ^ESCT1414545, ^ESCT1414546, ^ESCT1474197, ^ESCT1474201, ^ESCT1474299, ^ESCT1527612, ^ESCT1527615, ^ESCT1527616, ^ESCT1527618, ^ESCT1527620, ^ESCT1527621, ^ESCT1527622, ^ESCT1527624 | G301 |
| 2 | I | G30-4, G307, G3010, G304, G305, G306, G308, G30y0, G309, G3070, G30y1, G303, G30y2, G30X0, G3071, G300, G3011, G302, G30B, ^ESCTAC250300, ^ESCT1171766, ^ESCT1171768, ^ESCTAC334347, ^ESCTAC338160, ^ESCTAC345218, ^ESCTAC345956, ^ESCTAC355117, ^ESCTAC356587, ^ESCTAC364164, ^ESCTAC369992, ^ESCTAC374574, ^ESCTAC378491, ^ESCTAC477589, ^ESCTAC505629, ^ESCTAC505630, ^ESCTAC505631, ^ESCTAC505632, ^ESCTAC505633, ^ESCTAC505634, ^ESCTAC593532, ^ESCTNO604377, ^ESCTST665122, ^ESCTNS665139, ^ESCTAC757158, ^ESCTAC757159, ^ESCTAC757160, ^ESCTAC757161, ^ESCTAC757162, ^ESCTAC757232, ^ESCTAC757233, ^ESCTAC757234, ^ESCTAC757286, ^ESCTAC757288, ^ESCTAC757290, ^ESCTAC800426, ^ESCTAC801104, ^ESCTAC808775, ^ESCTAC984395, ^ESCTAC984396, ^ESCTAC984398, ^ESCTAC984399, ^ESCTAC984401, ^ESCTAC984402, ^ESCTAC984405, ^ESCTAC984407, ^ESCT1212233, ^ESCT1212234, ^ESCT1393516, ^ESCT1393519, ^ESCT1393520, ^ESCT1393521, ^ESCT1394109, ^ESCT1394111, ^ESCT1394114, ^ESCT1394116, ^ESCT1394121, ^ESCT1394123, ^ESCT1394521, ^ESCT1394523, ^ESCT1394772, ^ESCT1396130, ^ESCT1396131, ^ESCT1396134, ^ESCT1396135, ^ESCT1396137, ^ESCT1396138, ^ESCT1396142, ^ESCT1396143, ^ESCT1396144, ^ESCT1396146, ^ESCT1396147, ^ESCT1401503, ^ESCT1413294, ^ESCT1413295, ^ESCT1414537, ^ESCT1414539, ^ESCT1414543, ^ESCT1414545, ^ESCT1414546, ^ESCT1474197, ^ESCT1474201, ^ESCT1474299, ^ESCT1527612, ^ESCT1527615, ^ESCT1527616, ^ESCT1527618, ^ESCT1527621, ^ESCT1527624 | G301 |
| 3 | I | G30-4, G307, G3010, G304, G305, G306, G308, G30y0, G3070, G30y1, G303, G30y2, G30X0, G3071, G300, G3011, G302, G30B, ^ESCTAC250300, ^ESCT1171766, ^ESCT1171768, ^ESCTAC334347, ^ESCTAC338160, ^ESCTAC345218, ^ESCTAC345956, ^ESCTAC355117, ^ESCTAC356587, ^ESCTAC364164, ^ESCTAC369992, ^ESCTAC374574, ^ESCTAC378491, ^ESCTAC477589, ^ESCTAC505627, ^ESCTAC505629, ^ESCTAC505630, ^ESCTAC505631, ^ESCTAC505632, ^ESCTAC505633, ^ESCTAC505634, ^ESCTAC593532, ^ESCTNO604377, ^ESCTST665122, ^ESCTNS665139, ^ESCTAC757158, ^ESCTAC757159, ^ESCTAC757160, ^ESCTAC757161, ^ESCTAC757162, ^ESCTAC757232, ^ESCTAC757233, ^ESCTAC757234, ^ESCTAC757286, ^ESCTAC757288, ^ESCTAC757290, ^ESCTAC800426, ^ESCTAC801104, ^ESCTAC808775, ^ESCTAC984395, ^ESCTAC984396, ^ESCTAC984398, ^ESCTAC984399, ^ESCTAC984401, ^ESCTAC984402, ^ESCTAC984405, ^ESCTAC984407, ^ESCT1212233, ^ESCT1212234, ^ESCT1393516, ^ESCT1393519, ^ESCT1393520, ^ESCT1393521, ^ESCT1394109, ^ESCT1394111, ^ESCT1394114, ^ESCT1394116, ^ESCT1394121, ^ESCT1394123, ^ESCT1394521, ^ESCT1394523, ^ESCT1394598, ^ESCT1394759, ^ESCT1394772, ^ESCT1396130, ^ESCT1396131, ^ESCT1396134, ^ESCT1396135, ^ESCT1396137, ^ESCT1396138, ^ESCT1396142, ^ESCT1396143, ^ESCT1396144, ^ESCT1396145, ^ESCT1396146, ^ESCT1396147, ^ESCT1401503, ^ESCT1413294, ^ESCT1413295, ^ESCT1414537, ^ESCT1414539, ^ESCT1414543, ^ESCT1414545, ^ESCT1414546, ^ESCT1474197, ^ESCT1474201, ^ESCT1474299, ^ESCT1527612, ^ESCT1527615, ^ESCT1527616, ^ESCT1527618, ^ESCT1527620, ^ESCT1527621, ^ESCT1527622, ^ESCT1527624 | G301 |
| Wrist fracture - gpt-5.2 | | | |
| 1 | I | S241, S2351, S2341-1, S2347, S2349, S234B, S234C, S234D, S234E, S2351-1, S2359, S235A, S235B, S235C, S235E, S2400, S2409, S240A, S240B, S240C, S240D, S240E, S240F, S240z, S2410, S2419, S241A, S241B, S241C, S241D, S241E, S241F, S241y, S241z, S24z, S4C1, S23B, S23C, S2352, S235F, S234F, S2403, S2418, S2406, S2408, S2402, S2416, S2401, S2404, S2417, S2412, S2405, S2415, S2411, S2413, S2414, S2407, S2346, S2356, ^ESCT1167569, ^ESCT1167570, S2359-1, S2359-2, S2357, S4C36, S4C31, S4C1y, S4C15, S4C16, S4C11, S4C3, S4C3y, S4C14, S4C12, S235A-1, S235A-2, S234A, S234A-2, S234A-1, S4C04, S4C02, S4C0y, S4C05, S4C06, S4C01, S4C24, S4C22, S4C2y, S4C25, S4C26, S4C21, S2349-2, S2349-1, S2420, S2341-98, S234G, ^ESCTCL250422, ^ESCTOP262611, ^ESCTFR273650, ^ESCTFR301491, ^ESCTCL303424, ^ESCTOP305807, ^ESCTCL309869, ^ESCTOP321700, ^ESCTOP336361, ^ESCTBA338678, ^ESCTBA338679, ^ESCTOP359139, ^ESCTOP360729, ^ESCTFR371035, ^ESCTFR389651, ^ESCTOP393200, ^ESCTOP394930, ^ESCTAR421400, ^ESCTCL435380, ^ESCTOP435381, ^ESCTCO435824, ^ESCTRE435825, ^ESCTSM435826, ^ESCTCL484225, ^ESCTCL484228, ^ESCTCL484229, ^ESCTCL484892, ^ESCTCL484893, ^ESCTCL484896, ^ESCTCL484897, ^ESCTCL484898, ^ESCTOP484902, ^ESCTOP484905, ^ESCTOP484906, ^ESCTOP484907, ^ESCTCL484910, ^ESCTCL484913, ^ESCTCL484914, ^ESCTCL484915, ^ESCTOP484916, ^ESCTOP484921, ^ESCTFR546060, ^ESCTFR546062, ^ESCTFR546088, ^ESCTVO546201, ^ESCTVO546202, ^ESCTDO546203, ^ESCTDO546204, ^ESCTFR546212, ^ESCTTR565252, ^ESCTTR565254, ^ESCTTR565255, ^ESCTTR565256, ^ESCTFR565268, ^ESCTCL597198, ^ESCTPA730975, ^ESCTCL758786, ^ESCTPA766315, ^ESCT1221720, ^ESCT1370300, ^ESCT1370301, ^ESCT1379051, ^ESCT1419672, ^ESCT1507936, ^ESCT1507937, ^ESCT1565378, ^ESCTHU705948, ^ESCTCH705949 | S2341, S234, S235 |
| 2 | I | S241, S2351, S2341-1, S2347, S2349, S234B, S234C, S234D, S234E, S2351-1, S2359, S235A, S235B, S235C, S235E, S2400, S2409, S240A, S240B, S240C, S240D, S240E, S240F, S240z, S2410, S2419, S241A, S241B, S241C, S241D, S241E, S241F, S241y, S241z, S24z, S4C1, S23B, S23C, S2352, S235F, S234F, S2403, S2418, S2406, S2408, S2402, S2416, S2401, S2404, S2417, S2412, S2405, S2415, S2411, S2413, S2414, S2407, S2346, S2356, ^ESCT1167569, ^ESCT1167570, S2359-1, S2359-2, S2357, S4C36, S4C31, S4C1y, S4C15, S4C16, S4C11, S4C3, S4C3y, S4C14, S4C12, S235A-1, S235A-2, S234A, S234A-2, S234A-1, S4C04, S4C02, S4C0y, S4C05, S4C06, S4C01, S4C24, S4C22, S4C2y, S4C25, S4C26, S4C21, S2349-2, S2349-1, S2420, S2341-98, S234G, ^ESCTCL250422, ^ESCTOP262611, ^ESCTFR273650, ^ESCTFR301491, ^ESCTCL303424, ^ESCTOP305807, ^ESCTCL309869, ^ESCTOP321700, ^ESCTOP336361, ^ESCTBA338678, ^ESCTBA338679, ^ESCTOP359139, ^ESCTOP360729, ^ESCTFR371035, ^ESCTFR389651, ^ESCTOP393200, ^ESCTOP394930, ^ESCTAR421400, ^ESCTCL435380, ^ESCTOP435381, ^ESCTCO435824, ^ESCTRE435825, ^ESCTSM435826, ^ESCTCL484225, ^ESCTCL484228, ^ESCTCL484229, ^ESCTCL484892, ^ESCTCL484893, ^ESCTCL484896, ^ESCTCL484897, ^ESCTCL484898, ^ESCTOP484902, ^ESCTOP484905, ^ESCTOP484906, ^ESCTOP484907, ^ESCTCL484910, ^ESCTCL484913, ^ESCTCL484914, ^ESCTCL484915, ^ESCTOP484916, ^ESCTOP484921, ^ESCTFR546060, ^ESCTFR546062, ^ESCTFR546088, ^ESCTVO546201, ^ESCTVO546202, ^ESCTDO546203, ^ESCTDO546204, ^ESCTFR546212, ^ESCTTR565252, ^ESCTTR565254, ^ESCTTR565255, ^ESCTTR565256, ^ESCTFR565268, ^ESCTCL597198, ^ESCTPA730975, ^ESCTCL758786, ^ESCTPA766315, ^ESCT1221720, ^ESCT1370300, ^ESCT1370301, ^ESCT1379051, ^ESCT1419672, ^ESCT1507937, ^ESCT1565378, ^ESCTHU705948, ^ESCTCH705949 | S2341, S234, S235 |
| 3 | I | S2351, S2341-1, S2347, S2349, S234B, S234C, S234D, S2351-1, S2359, S235A, S235B, S235C, S235E, S2409, S240A, S240B, S240C, S240D, S240E, S2419, S241A, S241B, S241C, S241D, S241E, S241F, S241y, Syu63, S23B, S23C, S2341, S2352, S235F, S234F, S2418, S2406, S2408, S2402, S2416, S2401, S2404, S2417, S2412, S2405, S2415, S2411, S2413, S24-1, S2414, S2407, S2346, S2356, ^ESCT1167569, ^ESCT1167570, S2359-1, S2359-2, S2357, S4C36, S4C31, S4C1y, S4C15, S4C16, S4C11, S4C3, S4C3y, S4C14, S4C12, S235A-1, S235A-2, S234A, S234A-2, S234A-1, S4C04, S4C02, S4C0y, S4C05, S4C06, S4C01, S4C24, S4C22, S4C25, S4C26, S4C21, S2349-2, S2349-1, S2420, S2341-98, S2341-99, S234G, ^ESCTFR253074, ^ESCTFR253076, ^ESCTFR262070, ^ESCTFR262071, ^ESCTOP262611, ^ESCTFR273650, ^ESCTFR301489, ^ESCTFR301491, ^ESCTCL303424, ^ESCTOP305807, ^ESCTCL319032, ^ESCTOP321700, ^ESCTOP336361, ^ESCTBA338678, ^ESCTBA338679, ^ESCTOP359139, ^ESCTOP360729, ^ESCTFR369226, ^ESCTFR371035, ^ESCTFR386706, ^ESCTFR389651, ^ESCTOP393200, ^ESCTOP394930, ^ESCTAR421400, ^ESCTCL435380, ^ESCTOP435381, ^ESCTCO435824, ^ESCTRE435825, ^ESCTSM435826, ^ESCTCL484225, ^ESCTCL484228, ^ESCTCL484229, ^ESCTCL484892, ^ESCTCL484893, ^ESCTCL484896, ^ESCTCL484897, ^ESCTCL484898, ^ESCTOP484902, ^ESCTOP484905, ^ESCTOP484906, ^ESCTOP484907, ^ESCTCL484910, ^ESCTCL484913, ^ESCTCL484914, ^ESCTCL484915, ^ESCTOP484916, ^ESCTOP484921, ^ESCTFR546060, ^ESCTFR546062, ^ESCTFR546088, ^ESCTFR546199, ^ESCTVO546201, ^ESCTVO546202, ^ESCTDO546203, ^ESCTDO546204, ^ESCTFR546212, ^ESCTTR565252, ^ESCTTR565254, ^ESCTTR565255, ^ESCTTR565256, ^ESCTFR565268, ^ESCTCL597198, ^ESCTPA730975, ^ESCTCL758786, ^ESCTPA766315, ^ESCT1221720, ^ESCT1370303, ^ESCT1379051, ^ESCT1419672, ^ESCT1507936, ^ESCT1507937, ^ESCT1565378, ^ESCT1605454, ^ESCTHU705948, ^ESCTCH705949 | S241, S234, S235 |
| Vascular dementia - claude-sonnet-4-6 | | | |
| 1 | I | EMISICD10\|F0100, EMISICD10\|F0101, EMISICD10\|F0104, EMISICD10\|F0111, EMISICD10\|F0113, EMISICD10\|F0114, EMISICD10\|F0120, EMISICD10\|F0121, EMISICD10\|F0122, EMISICD10\|F0123, EMISICD10\|F0124, EMISICD10\|F0130, EMISICD10\|F0131, EMISICD10\|F0132, EMISICD10\|F0133, EMISICD10\|F0134, EMISICD10\|F0180, EMISICD10\|F0181, EMISICD10\|F0182, EMISICD10\|F0183, EMISICD10\|F0184, EMISICD10\|F0190, EMISICD10\|F0191, EMISICD10\|F0192, EMISICD10\|F0193, EMISICD10\|F0194, ^ESCT1480525 | F21y2 |
| 2 | I | EMISICD10\|F0100, EMISICD10\|F0101, EMISICD10\|F0104, EMISICD10\|F0111, EMISICD10\|F0113, EMISICD10\|F0114, EMISICD10\|F0120, EMISICD10\|F0121, EMISICD10\|F0122, EMISICD10\|F0123, EMISICD10\|F0124, EMISICD10\|F0130, EMISICD10\|F0131, EMISICD10\|F0132, EMISICD10\|F0133, EMISICD10\|F0134, EMISICD10\|F0180, EMISICD10\|F0181, EMISICD10\|F0182, EMISICD10\|F0183, EMISICD10\|F0184, EMISICD10\|F0190, EMISICD10\|F0191, EMISICD10\|F0192, EMISICD10\|F0193, EMISICD10\|F0194, ^ESCT1480525 |  |
| 3 | P | EMISICD10\|F0100, EMISICD10\|F0101, EMISICD10\|F0104, EMISICD10\|F0111, EMISICD10\|F0113, EMISICD10\|F0114, EMISICD10\|F0120, EMISICD10\|F0121, EMISICD10\|F0122, EMISICD10\|F0123, EMISICD10\|F0124, EMISICD10\|F0130, EMISICD10\|F0131, EMISICD10\|F0132, EMISICD10\|F0133, EMISICD10\|F0134, EMISICD10\|F0180, EMISICD10\|F0181, EMISICD10\|F0182, EMISICD10\|F0183, EMISICD10\|F0184, EMISICD10\|F0190, EMISICD10\|F0191, EMISICD10\|F0192, EMISICD10\|F0193, EMISICD10\|F0194 |  |
| Psoriasis - claude-sonnet-4-6 | | | |
| 1 | P | ^ESCTIN512406, ^ESCTVU666362 |  |
| 2 | P |  | ESCTFL1 |
| 3 | I | Nyu13, ESCTFL1, ^ESCTIN512406 |  |
| Hidradenitis suppurativa - claude-sonnet-4-6 | | | |
| 1 | P | ^ESCTVU667049 |  |
| 3 | I | ^ESCTVE346453, ^ESCTVU667049 | M25y1-1, EGTON66, ^ESCTSU346454, ^ESCTAX667047, ^ESCTAN667048, ^ESCT1501708 |
| Myocardial infarction - claude-sonnet-4-6 | | | |
| 1 | I | ^ESCTAC378491, ^ESCTAC505627, ^ESCTAC505629, ^ESCTAC505630, ^ESCTAC505631, ^ESCTAC505632, ^ESCTAC505633, ^ESCTAC505634, ^ESCTAC593532, ^ESCTNO604377, ^ESCTAC800426, ^ESCTAC801104, ^ESCTAC808775, ^ESCT1396145, ^ESCT1396146, ^ESCT1396147, ^ESCT1401503, ^ESCT1413294, ^ESCT1413295, ^ESCT1414537, ^ESCT1414539, ^ESCT1414543, ^ESCT1414545, ^ESCT1414546, ^ESCT1419470, ^ESCT1419471, ^ESCT1474197, ^ESCT1474201, ^ESCT1474299, ^ESCT1527612, ^ESCT1527615, ^ESCT1527616, ^ESCT1527618, ^ESCT1527620, ^ESCT1527621, ^ESCT1527622, ^ESCT1527624 | ^ESCTAC250300 |
| 2 | I | ^ESCT1171766, ^ESCTAC505627, ^ESCTAC505629, ^ESCTAC505630, ^ESCTAC505631, ^ESCTAC505632, ^ESCTAC505633, ^ESCTAC505634, ^ESCTNO604377, ^ESCTAC757159, ^ESCTAC808775, ^ESCT1401503, ^ESCT1413294, ^ESCT1413295, ^ESCT1419470, ^ESCT1419471, ^ESCT1474197, ^ESCT1474201, ^ESCT1474299, ^ESCT1527618, ^ESCT1527620, ^ESCT1527621, ^ESCT1527622, ^ESCT1527624 |  |
| 3 | I | Gyu34, G36, G30-99, ^ESCTMY285530, ^ESCTMI285534, ^ESCTMY285535, ^ESCTAM342720, ^ESCTAC356587, ^ESCTAC378491, ^ESCTNO604377 |  |
| Wrist fracture - claude-sonnet-4-6 | | | |
| 1 | I | S2409, S240A, S240B, S240C, S240D, S240E, S2419, S241A, S241B, S241C, S241D, S241E, S4C0, S4C1, Syu63, S2403, S2418, S2406, S2408, S2402, S2416, S2401, S2404, S2417, S2412, S2405, S2415, S2411, S2413, S2414, S2407, S4C36, S4C31, S4C1y, S4C15, S4C16, S4C11, S4C3, S4C3y, S4C14, S4C12, S4C04, S4C02, S4C0y, S4C05, S4C06, S4C01, S4C24, S4C22, S4C2, S4C2y, S4C25, S4C26, S4C21, S2420, S4C, S2341-98, S2341-99, ^ESCTCL250422, ^ESCTFR253074, ^ESCTFR253076, ^ESCTFR262068, ^ESCTFR262070, ^ESCTFR262071, ^ESCTOP262611, ^ESCTFR273648, ^ESCTFR273650, ^ESCTCL299359, ^ESCTFR301489, ^ESCTFR301491, ^ESCTOP305807, ^ESCTCL309869, ^ESCTCL310603, ^ESCTCL319032, ^ESCTCL320474, ^ESCTOP321700, ^ESCTOP336361, ^ESCTCL342815, ^ESCTOP359139, ^ESCTOP360729, ^ESCTFR369224, ^ESCTFR369226, ^ESCTFR371033, ^ESCTFR371035, ^ESCTFR386705, ^ESCTFR386706, ^ESCTFR389649, ^ESCTFR389651, ^ESCTOP393200, ^ESCTOP394930, ^ESCTCL396486, ^ESCTAR421400, ^ESCTCL484892, ^ESCTCL484893, ^ESCTCL484896, ^ESCTCL484897, ^ESCTCL484898, ^ESCTOP484902, ^ESCTOP484905, ^ESCTOP484906, ^ESCTOP484907, ^ESCTCL484910, ^ESCTCL484913, ^ESCTCL484914, ^ESCTCL484915, ^ESCTOP484916, ^ESCTOP484921, ^ESCTFR546059, ^ESCTFR546060, ^ESCTFR546062, ^ESCTFR546082, ^ESCTFR546088, ^ESCTTR565252, ^ESCTTR565254, ^ESCTTR565255, ^ESCTTR565256, ^ESCT1370300, ^ESCT1370301, ^ESCT1370303, ^ESCT1379051, ^ESCT1419672, ^ESCT1605454, ^ESCT1608054, ^ESCTHU705948, ^ESCTCH705949 | S241, S2351, S24, S2341-1, S2347, S2349, S234B, S234C, S234D, S234E, S2351-1, S2359, S235A, S235B, S235C, S235E, S2400, S240F, S240y, S240z, S2410, S241F, S241y, S241z, S24z, S23B, S23C, S2341, S2342, S2352, S235F, S234F, S234, S235, S240, S24-1, S2346, S2356, ^ESCT1167569, ^ESCT1167570, S2359-1, S2359-2, S2357, S235A-1, S235A-2, S234A, S234A-2, S234A-1, S2349-2, S2349-1, S234G, ^ESCTCL265096, ^ESCTCL277320, ^ESCTCL303424, ^ESCTBA338678, ^ESCTBA338679, ^ESCTFR383451, ^ESCTCL435380, ^ESCTOP435381, ^ESCTCO435824, ^ESCTRE435825, ^ESCTSM435826, ^ESCTCL484225, ^ESCTCL484228, ^ESCTCL484229, ^ESCTFR546199, ^ESCTVO546201, ^ESCTVO546202, ^ESCTDO546203, ^ESCTDO546204, ^ESCTFR546212, ^ESCTFR565268, ^ESCTCL597198, ^ESCTPA730975, ^ESCTCL758786, ^ESCTPA766315, ^ESCT1221720, ^ESCT1507936, ^ESCT1507937, ^ESCT1526232, ^ESCT1529546, ^ESCT1565378 |
| 2 | I | S2409, S240B, S240C, S240D, S240E, S240F, S2419, S241A, S241B, S241C, S241D, S241E, S241F, S4C0, S4C1, S2403, S2418, S2406, S2408, S2402, S2416, S2404, S2417, S2412, S2405, S2415, S2411, S2413, S2414, S2407, S4C36, S4C31, S4C1y, S4C15, S4C16, S4C11, S4C3, S4C3y, S4C14, S4C12, S4C04, S4C02, S4C0y, S4C05, S4C06, S4C01, S4C24, S4C22, S4C2, S4C2y, S4C25, S4C26, S4C21, S2420, S4C, S2341-99, ^ESCTCL250422, ^ESCTFR253074, ^ESCTFR253076, ^ESCTFR262068, ^ESCTFR262070, ^ESCTFR262071, ^ESCTOP262611, ^ESCTFR273648, ^ESCTFR273650, ^ESCTCL299359, ^ESCTFR301489, ^ESCTFR301491, ^ESCTOP305807, ^ESCTCL309869, ^ESCTCL310603, ^ESCTCL319032, ^ESCTCL320474, ^ESCTOP321700, ^ESCTOP336361, ^ESCTCL342815, ^ESCTOP359139, ^ESCTOP360729, ^ESCTFR369224, ^ESCTFR369226, ^ESCTFR371033, ^ESCTFR371035, ^ESCTFR386705, ^ESCTFR386706, ^ESCTFR389649, ^ESCTFR389651, ^ESCTOP393200, ^ESCTOP394930, ^ESCTCL396486, ^ESCTAR421400, ^ESCTCL484892, ^ESCTCL484893, ^ESCTCL484896, ^ESCTCL484897, ^ESCTCL484898, ^ESCTOP484902, ^ESCTOP484905, ^ESCTOP484906, ^ESCTOP484907, ^ESCTCL484910, ^ESCTCL484913, ^ESCTCL484914, ^ESCTCL484915, ^ESCTOP484916, ^ESCTOP484921, ^ESCTFR546059, ^ESCTFR546060, ^ESCTFR546062, ^ESCTFR546082, ^ESCTFR546088, ^ESCTTR565252, ^ESCTTR565254, ^ESCTTR565255, ^ESCTTR565256, ^ESCT1526232, ^ESCT1565378, ^ESCT1608054, ^ESCTHU705948, ^ESCTCH705949 | S241, S2351, S24, S2341-1, S2347, S2349, S234B, S234C, S234D, S234E, S2351-1, S2359, S235A, S235B, S235C, S235E, S2400, S240A, S240y, S240z, S2410, S241y, S241z, S24z, Syu63, S23B, S23C, S2341, S2342, S2352, S235F, S234F, S234, S235, S240, S2401, S24-1, S2346, S2356, ^ESCT1167569, ^ESCT1167570, S2359-1, S2359-2, S2357, S235A-1, S235A-2, S234A, S234A-2, S234A-1, S2349-2, S2349-1, S2341-98, S234G, ^ESCTCL265096, ^ESCTCL277320, ^ESCTCL303424, ^ESCTBA338678, ^ESCTBA338679, ^ESCTFR383451, ^ESCTCL435380, ^ESCTOP435381, ^ESCTCO435824, ^ESCTRE435825, ^ESCTSM435826, ^ESCTCL484225, ^ESCTCL484228, ^ESCTCL484229, ^ESCTFR546199, ^ESCTVO546201, ^ESCTVO546202, ^ESCTDO546203, ^ESCTDO546204, ^ESCTFR546212, ^ESCTFR565268, ^ESCTCL597198, ^ESCTPA730975, ^ESCTCL758786, ^ESCTPA766315, ^ESCT1221720, ^ESCT1370300, ^ESCT1370301, ^ESCT1370303, ^ESCT1379051, ^ESCT1419672, ^ESCT1507936, ^ESCT1507937, ^ESCT1529546, ^ESCT1605454 |
| 3 | I | S2409, S2419, S4C0, S4C1, S2418, S2406, S2408, S2416, S2417, S2415, S2407, S4C36, S4C31, S4C1y, S4C15, S4C16, S4C11, S4C3, S4C3y, S4C14, S4C12, S4C04, S4C02, S4C0y, S4C05, S4C06, S4C01, S4C24, S4C22, S4C2, S4C2y, S4C25, S4C26, S4C21, S4C, ^ESCTCL250422, ^ESCTFR273648, ^ESCTFR273650, ^ESCTOP305807, ^ESCTCL309869, ^ESCTOP321700, ^ESCTCL342815, ^ESCTOP359139, ^ESCTOP360729, ^ESCTFR369224, ^ESCTFR369226, ^ESCTFR371033, ^ESCTFR371035, ^ESCTFR389649, ^ESCTFR389651, ^ESCTOP393200, ^ESCTCL396486, ^ESCTAR421400, ^ESCTCL484892, ^ESCTCL484893, ^ESCTCL484896, ^ESCTCL484897, ^ESCTCL484898, ^ESCTOP484902, ^ESCTOP484905, ^ESCTOP484906, ^ESCTOP484907, ^ESCTCL484910, ^ESCTCL484913, ^ESCTCL484914, ^ESCTCL484915, ^ESCTOP484916, ^ESCTOP484921, ^ESCTFR546059, ^ESCTFR546060, ^ESCTFR546062, ^ESCTFR546082, ^ESCTFR546088, ^ESCTTR565252, ^ESCTTR565254, ^ESCTTR565255, ^ESCTTR565256, ^ESCTCL758786, ^ESCT1370300, ^ESCT1370301, ^ESCT1608054, ^ESCTHU705948, ^ESCTCH705949 | S241, S2351, S24, S2341-1, S2347, S2349, S234B, S234C, S234D, S234E, S2351-1, S2359, S235A, S235B, S235C, S235E, S2400, S240A, S240B, S240C, S240D, S240E, S240F, S240y, S240z, S2410, S241A, S241B, S241C, S241D, S241E, S241F, S241y, S241z, S24z, Syu63, S23B, S23C, S2341, S2342, S2352, S235F, S234F, S234, S235, S240, S2403, S2402, S2401, S2404, S2412, S2405, S2411, S2413, S24-1, S2414, S2346, S2356, ^ESCT1167569, ^ESCT1167570, S2359-1, S2359-2, S2357, S235A-1, S235A-2, S234A, S234A-2, S234A-1, S2349-2, S2349-1, S2420, S2341-98, S2341-99, S234G, ^ESCTFR253074, ^ESCTFR253076, ^ESCTFR262068, ^ESCTFR262070, ^ESCTFR262071, ^ESCTOP262611, ^ESCTCL277320, ^ESCTCL299359, ^ESCTFR301489, ^ESCTFR301491, ^ESCTCL303424, ^ESCTCL310603, ^ESCTCL319032, ^ESCTCL320474, ^ESCTOP336361, ^ESCTBA338678, ^ESCTBA338679, ^ESCTFR386705, ^ESCTFR386706, ^ESCTOP394930, ^ESCTCL435380, ^ESCTOP435381, ^ESCTCO435824, ^ESCTRE435825, ^ESCTSM435826, ^ESCTCL484225, ^ESCTCL484228, ^ESCTCL484229, ^ESCTFR546199, ^ESCTVO546201, ^ESCTVO546202, ^ESCTDO546203, ^ESCTDO546204, ^ESCTFR546212, ^ESCTFR565268, ^ESCTCL597198, ^ESCTPA730975, ^ESCTPA766315, ^ESCT1221720, ^ESCT1370303, ^ESCT1379051, ^ESCT1419672, ^ESCT1507936, ^ESCT1507937, ^ESCT1526232, ^ESCT1529546, ^ESCT1565378, ^ESCT1605454 |
| Atopic eczema - gemini-3-flash-preview | | | |
| 1 | I | M115 |  |
| 2 | I | M113, ^ESCTAT288430, ^ESCTAD288434, ^ESCTAT512285, ^ESCTAT512286, ^ESCTIN512287, ^ESCTDI512288, ^ESCTDI512289, ^ESCTDI512290, ^ESCTER512291, ^ESCTER512292, ^ESCTFO512293, ^ESCTFO512294, ^ESCTPR512295, ^ESCTPR512296, ^ESCTPH512297, ^ESCTPH512299, ^ESCTPH512300, ^ESCTPH512301, ^ESCTCH598917, ^ESCTCH598918, ^ESCTFL666116, ^ESCTFL666117, ^ESCTGE666120, ^ESCTGE666122, ^ESCTPR666124, ^ESCTXE666127, ^ESCTAT666130, ^ESCTAT666131, ^ESCTAT666132, ^ESCTAT666133, ^ESCTAT666134, ^ESCTAT666136, ^ESCTCH666138, ^ESCTIN666139, ^ESCTIN666140, ^ESCTCH666141, ^ESCTCH666142, ^ESCTAD666143, ^ESCTAD666144, ^ESCTAD666146, ^ESCTAD666148, ^ESCTIM666151, ^ESCTIN666152, ^ESCT1450246, ^ESCT1450704, ^ESCT1500750 |  |
| 3 | I | M115 |  |
| Vascular dementia - gemini-3-flash-preview | | | |
| 1 | I | E0042, EMISICD10\|F0100, EMISICD10\|F0101, EMISICD10\|F0104, EMISICD10\|F0111, EMISICD10\|F0113, EMISICD10\|F0114, EMISICD10\|F0120, EMISICD10\|F0121, EMISICD10\|F0122, EMISICD10\|F0123, EMISICD10\|F0124, EMISICD10\|F0130, EMISICD10\|F0131, EMISICD10\|F0132, EMISICD10\|F0133, EMISICD10\|F0134, EMISICD10\|F0180, EMISICD10\|F0181, EMISICD10\|F0182, EMISICD10\|F0183, EMISICD10\|F0184, EMISICD10\|F0190, EMISICD10\|F0191, EMISICD10\|F0192, EMISICD10\|F0193, EMISICD10\|F0194, ^ESCTMU272293, ^ESCTVA751506, ^ESCT1480525 |  |
| 2 | I | E0043, EMISICD10\|F0100, EMISICD10\|F0101, EMISICD10\|F0104, EMISICD10\|F0111, EMISICD10\|F0113, EMISICD10\|F0114, EMISICD10\|F0120, EMISICD10\|F0121, EMISICD10\|F0122, EMISICD10\|F0123, EMISICD10\|F0124, EMISICD10\|F0130, EMISICD10\|F0131, EMISICD10\|F0132, EMISICD10\|F0133, EMISICD10\|F0134, EMISICD10\|F0180, EMISICD10\|F0181, EMISICD10\|F0182, EMISICD10\|F0183, EMISICD10\|F0184, EMISICD10\|F0190, EMISICD10\|F0191, EMISICD10\|F0192, EMISICD10\|F0193, EMISICD10\|F0194, ^ESCTMU272293, ^ESCTVA751506, ^ESCTSU396465 |  |
| 3 | I | F21y2, E0041, E0042, E0043, F21y2-1, EMISICD10\|F0100, EMISICD10\|F0101, EMISICD10\|F0104, EMISICD10\|F0111, EMISICD10\|F0113, EMISICD10\|F0114, EMISICD10\|F0120, EMISICD10\|F0121, EMISICD10\|F0122, EMISICD10\|F0123, EMISICD10\|F0124, EMISICD10\|F0130, EMISICD10\|F0131, EMISICD10\|F0132, EMISICD10\|F0133, EMISICD10\|F0134, EMISICD10\|F0180, EMISICD10\|F0181, EMISICD10\|F0182, EMISICD10\|F0183, EMISICD10\|F0184, EMISICD10\|F0190, EMISICD10\|F0191, EMISICD10\|F0192, EMISICD10\|F0193, EMISICD10\|F0194, ^ESCTMU266460, ^ESCTVA266461, ^ESCTMU272293, ^ESCT1171774, ^ESCTMU291232, ^ESCTVA291233, ^ESCTMI341423, ^ESCTVA341425, ^ESCTMU341426, ^ESCTMU365367, ^ESCTSU396466, ^ESCTVA751506, ^ESCTIS784063, ^ESCTIS784064, ^ESCTMU803324, ^ESCTVA808923, ^ESCTVA808924, ^ESCTVA990225, ^ESCT1393438, ^ESCT1480525, ^ESCTSU396465 | E0040, E004z, Eu01y, Eu01z, E004, Eu01-1, Eu011, Eu012, Eu01, Eu010, E004-1, EMISNQDV1 |
| Eosinophilic esophagitis - gemini-3-flash-preview | | | |
| 1 | I | J1017, ^ESCT1165474, ^ESCTEO508265 |  |
| 2 | C | ^ESCT1165474, ^ESCTEO508265 |  |
| Psoriasis - gemini-3-flash-preview | | | |
| 1 | I | M1612, Myu30, M161J, ^ESCTAC255528, ^ESCTLO382192, ^ESCTEC512389, ^ESCTEC512390, ^ESCTPS512393, ^ESCTPS512395, ^ESCTGE512402, ^ESCTCH666357, ^ESCTGU666360, ^ESCTHY666374, ^ESCTHY666375, ^ESCTAC666380, ^ESCTCH666382, ^ESCT1363707 | ESCTFL1 |
| 2 | I | ^ESCTPS512393, ^ESCTPS512395 |  |
| 3 | I | M166, ^ESCTPS512392, ^ESCTPS512393, ^ESCTPS512395, ^ESCTPE666366 |  |
| Hidradenitis suppurativa - gemini-3-flash-preview | | | |
| 2 | I | ^ESCTVE346453 |  |
| Myocardial infarction - gemini-3-flash-preview | | | |
| 1 | I | G30-4, G3010, G304, G305, G306, G30y0, G309, G30y1, G303, G30y2, G30X0, Gyu34, G300, G3011, G302, G30B, ^ESCTAC250300, ^ESCT1171768, ^ESCTMY285535, ^ESCTAC334347, ^ESCTAC338160, ^ESCTAC345218, ^ESCTAC345956, ^ESCTAC355117, ^ESCTAC356587, ^ESCTAC364164, ^ESCTAC369992, ^ESCTAC378491, ^ESCTAC505627, ^ESCTAC505629, ^ESCTAC505631, ^ESCTAC505633, ^ESCTAC593532, ^ESCTRE709309, ^ESCTAC757158, ^ESCTAC757159, ^ESCTAC757161, ^ESCTAC757162, ^ESCTAC757232, ^ESCTAC757234, ^ESCTAC757286, ^ESCTAC757288, ^ESCTAC800426, ^ESCTAC801104, ^ESCTAC808775, ^ESCTAC984395, ^ESCTAC984396, ^ESCTAC984398, ^ESCTAC984399, ^ESCTAC984401, ^ESCTAC984402, ^ESCTAC984405, ^ESCTAC984407, ^ESCT1212233, ^ESCT1212234, ^ESCT1393516, ^ESCT1393520, ^ESCT1393521, ^ESCT1394109, ^ESCT1394111, ^ESCT1394114, ^ESCT1394116, ^ESCT1394121, ^ESCT1394123, ^ESCT1394521, ^ESCT1394523, ^ESCT1394598, ^ESCT1394759, ^ESCT1394772, ^ESCT1396130, ^ESCT1396131, ^ESCT1396134, ^ESCT1396135, ^ESCT1396137, ^ESCT1396142, ^ESCT1396144, ^ESCT1396146, ^ESCT1414537, ^ESCT1414539, ^ESCT1414543, ^ESCT1414546, ^ESCT1527612, ^ESCT1527615, ^ESCT1527618, ^ESCT1527621, ^ESCT1527624 | G301 |
| 2 | I | G303, G302, ^ESCTAC334347, ^ESCTAC345956, ^ESCTAC355117, ^ESCTAC356587, ^ESCTAC374574, ^ESCTAC800426, ^ESCTAC801104, ^ESCT1393519, ^ESCT1394772, ^ESCT1396135, ^ESCT1414537, ^ESCT1414543 |  |
| 3 | I | G3010, G306, G308, G303, Gyu34, G302, ^ESCTAC250300, ^ESCT1171766, ^ESCTAC334347, ^ESCTAC345218, ^ESCTAC355117, ^ESCTAC356587, ^ESCTAC364164, ^ESCTAC374574, ^ESCTAC378491, ^ESCTAC505627, ^ESCTAC505629, ^ESCTAC505630, ^ESCTAC505631, ^ESCTAC505632, ^ESCTAC505633, ^ESCTAC505634, ^ESCTNO604377, ^ESCTAC757158, ^ESCTAC757159, ^ESCTAC757160, ^ESCTAC757161, ^ESCTAC757162, ^ESCTAC757232, ^ESCTAC757233, ^ESCTAC757234, ^ESCTAC757286, ^ESCTAC757288, ^ESCTAC757290, ^ESCTAC800426, ^ESCTAC801104, ^ESCTAC808775, ^ESCTAC984395, ^ESCTAC984396, ^ESCTAC984398, ^ESCTAC984399, ^ESCTAC984401, ^ESCTAC984402, ^ESCTAC984405, ^ESCTAC984407, ^ESCT1212233, ^ESCT1212234, ^ESCT1393516, ^ESCT1393519, ^ESCT1393521, ^ESCT1394109, ^ESCT1394111, ^ESCT1394114, ^ESCT1394116, ^ESCT1394121, ^ESCT1394123, ^ESCT1394521, ^ESCT1394523, ^ESCT1394598, ^ESCT1394759, ^ESCT1394772, ^ESCT1396130, ^ESCT1396131, ^ESCT1396134, ^ESCT1396135, ^ESCT1396137, ^ESCT1396138, ^ESCT1396142, ^ESCT1396143, ^ESCT1396144, ^ESCT1396145, ^ESCT1396146, ^ESCT1396147, ^ESCT1413294, ^ESCT1413295, ^ESCT1414537, ^ESCT1414539, ^ESCT1414543, ^ESCT1414545, ^ESCT1414546, ^ESCT1474197, ^ESCT1474201, ^ESCT1474299, ^ESCT1527612, ^ESCT1527615, ^ESCT1527616, ^ESCT1527618, ^ESCT1527620, ^ESCT1527621, ^ESCT1527622, ^ESCT1527624 |  |
| Wrist fracture - gemini-3-flash-preview | | | |
| 1 | I | S241, S234C, S234D, S234E, S235C, S235E, S2400, S2409, S240A, S240B, S240C, S240D, S240E, S240F, S240y, S240z, S2410, S2419, S241A, S241B, S241C, S241D, S241E, S241F, S241y, S241z, S24z, S4C1, Syu63, S23B, S23C, S2352, S2403, S2418, S2406, S2408, S2416, S2401, S2404, S2417, S2412, S2405, S2415, S2411, S2413, S2414, S2407, S2346, S2356, ^ESCT1167569, ^ESCT1167570, S2359-1, S2359-2, S4C36, S4C31, S4C1y, S4C15, S4C16, S4C11, S4C3, S4C3y, S4C14, S4C12, S235A-1, S235A-2, S234A-2, S234A-1, S4C04, S4C02, S4C0y, S4C05, S4C06, S4C01, S4C24, S4C22, S4C2y, S4C25, S4C26, S4C21, S2349-2, S2349-1, S2341-98, S234G, ^ESCTCL250422, ^ESCTFR253076, ^ESCTFR262070, ^ESCTFR262071, ^ESCTOP262611, ^ESCTCL265096, ^ESCTFR273650, ^ESCTFR301489, ^ESCTFR301491, ^ESCTCL303424, ^ESCTOP305807, ^ESCTCL309869, ^ESCTOP321700, ^ESCTOP336361, ^ESCTBA338679, ^ESCTOP359139, ^ESCTOP360729, ^ESCTFR369226, ^ESCTFR371035, ^ESCTFR383451, ^ESCTFR386706, ^ESCTFR389651, ^ESCTOP393200, ^ESCTOP394930, ^ESCTAR421400, ^ESCTCL435380, ^ESCTOP435381, ^ESCTRE435825, ^ESCTCL484225, ^ESCTCL484228, ^ESCTCL484229, ^ESCTCL484892, ^ESCTCL484893, ^ESCTCL484896, ^ESCTCL484897, ^ESCTCL484898, ^ESCTOP484902, ^ESCTOP484905, ^ESCTOP484906, ^ESCTOP484907, ^ESCTCL484910, ^ESCTCL484913, ^ESCTCL484914, ^ESCTCL484915, ^ESCTOP484916, ^ESCTOP484921, ^ESCTFR546060, ^ESCTFR546062, ^ESCTFR546088, ^ESCTVO546202, ^ESCTDO546204, ^ESCTFR546212, ^ESCTTR565252, ^ESCTTR565254, ^ESCTTR565255, ^ESCTTR565256, ^ESCTPA730975, ^ESCTCL758786, ^ESCTPA766315, ^ESCT1370300, ^ESCT1370301, ^ESCT1370303, ^ESCT1379051, ^ESCT1507936, ^ESCT1507937, ^ESCT1526232, ^ESCT1529546, ^ESCT1565378, ^ESCT1608054, ^ESCTHU705948 | S2351, S2341, S234, S235 |
| 2 | I | S234C, S234D, S234E, S235C, S235E, S2400, S2409, S240D, S240E, S240F, S240y, S240z, S2410, S2419, S241A, S241B, S241C, S241D, S241E, S241F, S241y, S241z, S24z, S4C1, Syu63, S23B, S23C, S2352, S2403, S2418, S2406, S2408, S2416, S2417, S2405, S2415, S2413, S24-1, S2414, ^ESCT1167569, ^ESCT1167570, S2359-1, S2359-2, S4C36, S4C31, S4C1y, S4C15, S4C16, S4C11, S4C3, S4C3y, S4C14, S4C12, S235A-1, S235A-2, S234A-2, S234A-1, S4C04, S4C02, S4C0y, S4C05, S4C01, S4C24, S4C22, S4C2y, S4C25, S4C26, S4C21, S2349-2, S2349-1, S2341-98, S234G, ^ESCTCL250422, ^ESCTFR253076, ^ESCTFR262070, ^ESCTFR262071, ^ESCTOP262611, ^ESCTCL265096, ^ESCTFR273650, ^ESCTCL277320, ^ESCTCL299359, ^ESCTFR301489, ^ESCTFR301491, ^ESCTCL303424, ^ESCTOP305807, ^ESCTCL309869, ^ESCTCL310603, ^ESCTCL319032, ^ESCTCL320474, ^ESCTOP321700, ^ESCTOP336361, ^ESCTBA338679, ^ESCTCL342815, ^ESCTOP359139, ^ESCTOP360729, ^ESCTFR369226, ^ESCTFR371035, ^ESCTFR386706, ^ESCTFR389651, ^ESCTOP393200, ^ESCTOP394930, ^ESCTCL396486, ^ESCTAR421400, ^ESCTCL435380, ^ESCTOP435381, ^ESCTCO435824, ^ESCTRE435825, ^ESCTCL484225, ^ESCTCL484228, ^ESCTCL484229, ^ESCTCL484892, ^ESCTCL484893, ^ESCTCL484896, ^ESCTCL484897, ^ESCTCL484898, ^ESCTOP484902, ^ESCTOP484905, ^ESCTOP484906, ^ESCTOP484907, ^ESCTCL484910, ^ESCTCL484913, ^ESCTCL484914, ^ESCTCL484915, ^ESCTOP484916, ^ESCTOP484921, ^ESCTFR546062, ^ESCTFR546088, ^ESCTFR546199, ^ESCTVO546201, ^ESCTVO546202, ^ESCTDO546203, ^ESCTDO546204, ^ESCTFR546212, ^ESCTTR565252, ^ESCTTR565254, ^ESCTTR565255, ^ESCTTR565256, ^ESCTCL597198, ^ESCTPA730975, ^ESCTCL758786, ^ESCTPA766315, ^ESCT1221720, ^ESCT1370300, ^ESCT1370301, ^ESCT1370303, ^ESCT1379051, ^ESCT1419672, ^ESCT1507936, ^ESCT1507937, ^ESCT1526232, ^ESCT1529546, ^ESCT1565378, ^ESCT1605454, ^ESCT1608054, ^ESCTHU705948 | S241, S2351, S2341, S234, S235, S240 |
| 3 | I | S234D, S235B, S235C, S235E, S2419, S23B, S23C, S2352, S2418, S2406, S2416, S2404, S2417, S2415, S2413, S2414, S2356, ^ESCT1167569, ^ESCT1167570, S2359-1, S2359-2, S234-1, S4C36, S4C31, S4C1y, S4C15, S4C16, S4C11, S4C3, S4C3y, S4C14, S4C12, S235A-1, S235A-2, S234A-2, S234A-1, S4C04, S4C02, S4C0y, S4C05, S4C22, S4C2y, S4C25, S4C21, S2349-2, S2349-1, S234G, ^ESCTCL250422, ^ESCTFR253076, ^ESCTFR262070, ^ESCTFR262071, ^ESCTOP262611, ^ESCTFR273650, ^ESCTCL303424, ^ESCTOP305807, ^ESCTOP321700, ^ESCTOP336361, ^ESCTBA338679, ^ESCTOP359139, ^ESCTOP360729, ^ESCTFR369224, ^ESCTFR369226, ^ESCTFR371033, ^ESCTFR371035, ^ESCTFR383451, ^ESCTFR386706, ^ESCTFR389651, ^ESCTOP393200, ^ESCTOP394930, ^ESCTAR421400, ^ESCTOP435381, ^ESCTRE435825, ^ESCTCL484893, ^ESCTCL484897, ^ESCTOP484902, ^ESCTOP484905, ^ESCTOP484906, ^ESCTOP484907, ^ESCTCL484910, ^ESCTCL484913, ^ESCTCL484914, ^ESCTCL484915, ^ESCTOP484916, ^ESCTOP484921, ^ESCTFR546060, ^ESCTFR546062, ^ESCTFR546088, ^ESCTVO546201, ^ESCTVO546202, ^ESCTDO546203, ^ESCTDO546204, ^ESCTTR565252, ^ESCTTR565254, ^ESCTTR565255, ^ESCTTR565256, ^ESCTFR565268, ^ESCTPA730975, ^ESCTPA766315, ^ESCT1221720, ^ESCT1370300, ^ESCT1370301, ^ESCT1370303, ^ESCT1379051, ^ESCT1419672, ^ESCT1507936, ^ESCT1507937, ^ESCT1526232, ^ESCT1529546, ^ESCT1565378, ^ESCTHU705948 | S235 |
