## Supplementary Tables for "Large language models and retrieval augmented generation for complex clinical codelists: evaluating performance and assessing failure modes": Supplementary Table 3.html

| Epoch | Score | Required | Retrieved | Not retrieved | Unused | % retrieved |
| --- | --- | --- | --- | --- | --- | --- |
| Atopic eczema - gemini-3-pro-preview | | | | | | |
| --- | --- | --- | --- | --- | --- | --- |
| 1 | I | 56 | 54 | 2 | 0 | 96% |
| 2 | I | 56 | 47 | 9 | 0 | 84% |
| 3 | C | 56 | 56 | 0 | 0 | 100% |
| Vascular dementia - gemini-3-pro-preview | | | | | | |
| 1 | I | 63 | 36 | 27 | 0 | 57% |
| 2 | C | 63 | 37 | 26 | 0 | 59% |
| 3 | C | 63 | 37 | 26 | 0 | 59% |
| Eosinophilic esophagitis - gemini-3-pro-preview | | | | | | |
| 1 | C | 3 | 3 | 0 | 0 | 100% |
| 2 | C | 3 | 3 | 0 | 0 | 100% |
| 3 | C | 3 | 3 | 0 | 0 | 100% |
| Psoriasis - gemini-3-pro-preview | | | | | | |
| 1 | I | 76 | 74 | 2 | 0 | 97% |
| 2 | I | 76 | 74 | 2 | 0 | 97% |
| 3 | I | 76 | 74 | 2 | 0 | 97% |
| Hidradenitis suppurativa - gemini-3-pro-preview | | | | | | |
| 1 | C | 8 | 8 | 0 | 0 | 100% |
| 2 | C | 8 | 8 | 0 | 0 | 100% |
| 3 | C | 8 | 8 | 0 | 0 | 100% |
| Myocardial infarction - gemini-3-pro-preview | | | | | | |
| 1 | I | 132 | 70 | 62 | 0 | 53% |
| 2 | I | 132 | 113 | 19 | 0 | 86% |
| 3 | I | 132 | 102 | 30 | 0 | 77% |
| Wrist fracture - gemini-3-pro-preview | | | | | | |
| 1 | I | 214 | 74 | 140 | 2 | 35% |
| 2 | I | 214 | 104 | 110 | 3 | 49% |
| 3 | I | 214 | 123 | 91 | 0 | 57% |
| Atopic eczema - gpt-5.2 | | | | | | |
| 1 | I | 56 | 48 | 8 | 0 | 86% |
| 2 | I | 56 | 52 | 4 | 0 | 93% |
| 3 | I | 56 | 48 | 8 | 0 | 86% |
| Vascular dementia - gpt-5.2 | | | | | | |
| 1 | I | 64 | 26 | 38 | 0 | 41% |
| 2 | I | 64 | 26 | 38 | 0 | 41% |
| 3 | I | 64 | 25 | 39 | 25 | 39% |
| Eosinophilic esophagitis - gpt-5.2 | | | | | | |
| 1 | C | 3 | 3 | 0 | 0 | 100% |
| 2 | C | 3 | 3 | 0 | 0 | 100% |
| 3 | C | 3 | 3 | 0 | 0 | 100% |
| Psoriasis - gpt-5.2 | | | | | | |
| 1 | I | 78 | 63 | 15 | 1 | 81% |
| 2 | I | 78 | 40 | 38 | 0 | 51% |
| 3 | I | 78 | 52 | 26 | 0 | 67% |
| Hidradenitis suppurativa - gpt-5.2 | | | | | | |
| 1 | I | 8 | 7 | 1 | 0 | 88% |
| 2 | I | 8 | 7 | 1 | 0 | 88% |
| 3 | I | 8 | 7 | 1 | 0 | 88% |
| Myocardial infarction - gpt-5.2 | | | | | | |
| 1 | I | 132 | 17 | 115 | 1 | 13% |
| 2 | I | 132 | 24 | 108 | 1 | 18% |
| 3 | I | 132 | 19 | 113 | 1 | 14% |
| Wrist fracture - gpt-5.2 | | | | | | |
| 1 | I | 214 | 50 | 164 | 3 | 23% |
| 2 | I | 214 | 51 | 163 | 3 | 24% |
| 3 | I | 214 | 50 | 164 | 3 | 23% |
| Atopic eczema - claude-sonnet-4-6 | | | | | | |
| 1 | P | 56 | 56 | 0 | 0 | 100% |
| 2 | P | 56 | 56 | 0 | 0 | 100% |
| 3 | P | 56 | 56 | 0 | 0 | 100% |
| Vascular dementia - claude-sonnet-4-6 | | | | | | |
| 1 | I | 64 | 37 | 27 | 1 | 58% |
| 2 | I | 64 | 37 | 27 | 0 | 58% |
| 3 | P | 64 | 38 | 26 | 0 | 59% |
| Eosinophilic esophagitis - claude-sonnet-4-6 | | | | | | |
| 1 | C | 3 | 3 | 0 | 0 | 100% |
| 2 | C | 3 | 3 | 0 | 0 | 100% |
| 3 | C | 3 | 3 | 0 | 0 | 100% |
| Psoriasis - claude-sonnet-4-6 | | | | | | |
| 1 | P | 76 | 74 | 2 | 0 | 97% |
| 2 | P | 76 | 76 | 0 | 1 | 100% |
| 3 | I | 76 | 73 | 3 | 0 | 96% |
| Hidradenitis suppurativa - claude-sonnet-4-6 | | | | | | |
| 1 | P | 8 | 7 | 1 | 0 | 88% |
| 2 | C | 8 | 8 | 0 | 0 | 100% |
| 3 | I | 8 | 6 | 2 | 6 | 75% |
| Myocardial infarction - claude-sonnet-4-6 | | | | | | |
| 1 | I | 132 | 95 | 37 | 1 | 72% |
| 2 | I | 132 | 108 | 24 | 0 | 82% |
| 3 | I | 132 | 122 | 10 | 0 | 92% |
| Wrist fracture - claude-sonnet-4-6 | | | | | | |
| 1 | I | 214 | 89 | 125 | 82 | 42% |
| 2 | I | 214 | 95 | 119 | 88 | 44% |
| 3 | I | 214 | 131 | 83 | 122 | 61% |
| Atopic eczema - gemini-3-flash-preview | | | | | | |
| 1 | I | 56 | 55 | 1 | 0 | 98% |
| 2 | I | 56 | 9 | 47 | 0 | 16% |
| 3 | I | 56 | 55 | 1 | 0 | 98% |
| Vascular dementia - gemini-3-flash-preview | | | | | | |
| 1 | I | 64 | 34 | 30 | 0 | 53% |
| 2 | I | 64 | 34 | 30 | 0 | 53% |
| 3 | I | 64 | 12 | 52 | 12 | 19% |
| Eosinophilic esophagitis - gemini-3-flash-preview | | | | | | |
| 1 | I | 3 | 0 | 3 | 0 | 0% |
| 2 | C | 3 | 1 | 2 | 0 | 33% |
| 3 | C | 3 | 3 | 0 | 0 | 100% |
| Psoriasis - gemini-3-flash-preview | | | | | | |
| 1 | I | 76 | 59 | 17 | 1 | 78% |
| 2 | I | 76 | 74 | 2 | 0 | 97% |
| 3 | I | 76 | 71 | 5 | 0 | 93% |
| Hidradenitis suppurativa - gemini-3-flash-preview | | | | | | |
| 1 | C | 8 | 8 | 0 | 0 | 100% |
| 2 | I | 8 | 7 | 1 | 0 | 88% |
| 3 | C | 8 | 8 | 0 | 0 | 100% |
| Myocardial infarction - gemini-3-flash-preview | | | | | | |
| 1 | I | 132 | 46 | 86 | 1 | 35% |
| 2 | I | 132 | 118 | 14 | 0 | 89% |
| 3 | I | 132 | 41 | 91 | 0 | 31% |
| Wrist fracture - gemini-3-flash-preview | | | | | | |
| 1 | I | 214 | 61 | 153 | 4 | 29% |
| 2 | I | 214 | 59 | 155 | 6 | 28% |
| 3 | I | 214 | 104 | 110 | 1 | 49% |
