## Supplementary Tables for "Large language models and retrieval augmented generation for complex clinical codelists: evaluating performance and assessing failure modes": Supplementary Table 4.html

| codelist | summary |
| --- | --- |
| Atopic eczema | Claude Sonnet 4.6 demonstrated high accuracy across most runs but missed ^ESCTER512291 **Erythrodermic atopic dermatitis** and ^ESCTER512292 **Erythrodermic atopic eczema** in one epoch due to character substitution errors. Gemini 3 Flash Preview initially failed to include specific clinical patterns, missing ^ESCTIN512287 **Inverse pattern atopic dermatitis**, ^ESCTDI512288 **Discoid atopic dermatitis**, and ^ESCTPR666124 **Prurigo pattern atopic dermatitis**. GPT 5.2 consistently struggled with standard Read codes across all epochs, frequently omitting M114 **Allergic (intrinsic) eczema**, M112 **Infantile eczema**, M113 **Flexural eczema**, and M115 **Besnier’s prurigo**. |
| Eosinophilic esophagitis | Across all epochs, no required codes were missed by claude-sonnet-4, gemini-3-flash, or gpt-5.2. Every model consistently included all mandatory codes, including J1017 **Eosinophilic oesophagitis**, ^ESCT1165474 **Food-induced eosinophilic oesophagitis**, and ^ESCTEO508265 **Eosinophilic esophagitis**. There were no differences between models regarding required code coverage. |
| Hidradenitis suppurativa | Claude-sonnet-4-6 consistently missed EGTON66 **Hydradenitis suppurativa** and often omitted ^ESCTVE346453 **Verneuil’s disease**. Gemini-3-flash-preview was the only model to include all required codes in one epoch, though it also missed EGTON66 **Hydradenitis suppurativa** and ^ESCTVE346453 **Verneuil’s disease** in other runs. GPT-5.2 exhibited the highest consistency, missing only EGTON66 **Hydradenitis suppurativa** across all three epochs. |
| Myocardial infarction | Claude-sonnet consistently omitted core codes such as ^ESCTMY285535 **Myocardial infarct** and Q-wave specific codes like ^ESCTAC505627 **Acute Q wave infarction - anteroseptal**. Gemini-3-flash failed to capture broad SNOMED definitions including ^ESCTMY285530 **Myocardial infarction** and ^ESCTMI285534 **MI - Myocardial infarction**, as well as arterial occlusion details like ^ESCTAC801104 **Acute myocardial infarction due to right coronary artery occlusion**. GPT-5.2 was distinguished by its omission of Read codes such as G3010 **Acute anteroapical infarction** and Gyu34 **[X]Acute transmural myocardial infarction of unspecif site**, alongside anatomical SNOMED markers like ^ESCTAC250300 **Acute myocardial infarction of basal-lateral wall**. Common omissions across models included G36 **Myocardial infarction with complication** and specialized artery occlusion codes like ^ESCT1394109 **Acute ST segment elevation myocardial infarction due to proximal left anterior descending coronary artery occlusion**. |
| Psoriasis | Claude-Sonnet-4-6 repeatedly failed to identify required alphanumeric codes, notably missing ^ESCTAC255528 **Acute palmoplantar pustular psoriasis**, ^ESCTCH294145 **Chronic palmoplantar pustular psoriasis**, and ^ESCTGE311787 **Generalised pustular psoriasis of von Zumbush**, along with nail-specific codes such as ^ESCTPS512392 **Psoriatic nail dystrophy** and ^ESCTPS512395 **Psoriatic onycholysis**. GPT-5.2 consistently omitted the nail-related codes ^ESCTPS512393 **Psoriatic nail involvement** and ^ESCTPS512394 **Psoriatic nail pitting**, while also failing to include specific clinical presentations like ^ESCTGE666367 **Generalised psoriasis**, ^ESCTAC666380 **Acute generalised pustular flare of preexisting plaque psoriasis**, and ^ESCTCH666382 **Childhood pustular psoriasis**. In contrast, Gemini-3-flash-preview successfully retrieved all required codes across all epochs, exhibiting higher sensitivity to the complex ^ESCT terminology than the other models. |
| Vascular dementia | Claude-sonnet-4-6 consistently missed F21y2 **Binswanger’s disease**, F21y2-1 **Binswanger’s encephalopathy**, ^ESCTMU341426 **Multi infarct dementia**, and ^ESCTSU396465 **Subcortical arteriosclerotic encephalopathy**. Gemini-3-flash-preview also missed ^ESCTMU341426 **Multi infarct dementia** and ^ESCTSU396465 **Subcortical arteriosclerotic encephalopathy**, but was unique in omitting ^ESCTVA751506 **Vascular dementia in remission** and ^ESCT1480525 **Behavioural disturbance due to multi-infarct dementia**. GPT-5.2 missed the widest range of codes, including the Binswanger’s terms and the E004 series such as E004 **Arteriosclerotic dementia**, E0040 **Uncomplicated arteriosclerotic dementia**, and E0041 **Arteriosclerotic dementia with delirium**, alongside ^ESCTSU396466 **Subcortical atherosclerotic dementia**. |
| Wrist fracture | Claude-sonnet omitted high-level codes like S23B **Fracture of lower end of radius** and S24 **Fracture of carpal bone**, alongside mapping codes such as ^ESCT1167569 **Fracture of lower end of radius with dorsal tilt**. Gemini-3-flash-preview lacked specific intra-articular and dislocation codes, for example S235C **Open fracture distal radius, intra-articular, die-punch** and S4C1y **Open fracture-dislocation other carpal**. Gpt-5.2 failed to list specific carpal bones such as S2403 **Closed fracture triquetral** and S2413 **Open fracture triquetral**, and specifically excluded ^ESCT1379051 **Disorder due to and following fracture at wrist and/or hand level**. All models missed Syu63 **[X]Fracture of other carpal bone(s)**. |
