## Supplementary Tables for "Large language models and retrieval augmented generation for complex clinical codelists: evaluating performance and assessing failure modes": Supplementary Table 5.html

| codelist | summary |
| --- | --- |
| Atopic eczema | Claude-sonnet and Gemini-3-flash consistently included optional codes such as M116 **Neurodermatitis - diffuse**, M116-1 **Brocq’s neurodermatitis**, ^ESCTAL288431 **Allergic eczema**, and ^ESCTEN512284 **Endogenous eczema**. Claude-sonnet uniquely introduced invalid identifiers such as ^ESCTEM512291 **Erythrodermic atopic dermatitis**. Gemini-3-flash included broader terms like M12z1 **Eczema**, ^ESCTGE666346 **Generalised eczema**, and numerous contact dermatitis codes, including ^ESCTCO512283 **Contact dermatitis** and ^ESCT1379223 **Allergic contact dermatitis**. GPT-5.2 was more conservative, only including a few optional codes such as M1831 **Pompholyx** and M116-99 **Neurodermatitis**. |
| Eosinophilic esophagitis | Across all evaluation epochs, Claude Sonnet, Gemini Flash Preview, and GPT-5.2 did not include any irrelevant codes outside of the target codelist. All models correctly restricted their selections to codes present in the criterion, such as J1017 **Eosinophilic oesophagitis**, ^ESCT1165474 **Food-induced eosinophilic oesophagitis**, and ^ESCTEO508265 **Eosinophilic esophagitis**. The optional code ^ESCTEO508286 **Eosinophilic ulcer of oesophagus** was occasionally included by Claude Sonnet and Gemini Flash Preview, but it remained within the acceptable target list. |
| Hidradenitis suppurativa | Across all submissions and models, including Claude-sonnet-4-6, Gemini-3-flash-preview, and GPT-5.2, no irrelevant codes were included. All codes provided in the submissions were verified against the target codelist as either required or optional. |
| Myocardial infarction | Claude-sonnet-4-6 frequently included irrelevant codes for past events and complications, such as G32 **Old myocardial infarction**, G32-1 **Old myocardial infarction**, G32-2 **Old myocardial infarction**, G35 **Subsequent myocardial infarction**, and G38 **Postoperative myocardial infarction**. It also introduced history-based codes like ^ESCTHI751033 **History of non-ST segment elevation myocardial infarction** and ^ESCTOL505641 **Old myocardial infarction**. GPT-5.2 showed similar patterns, including the G32 series and 14AT **History of myocardial infarction**. Gemini-3-flash-preview was the most selective, only erroneously including G32 **Old myocardial infarction** in one instance. |
| Psoriasis | claude-sonnet-4-6 and gemini-3-flash-preview both frequently included irrelevant codes for parapsoriasis and specific inflammatory conditions. claude-sonnet-4-6 included M162 Parapsoriasis, Myu31 Parapsoriasis, M1602 Arthritis mutilans, M161G Acrodermatitis continua, and ^ESCTAC386403 Acrodermatitis continua of Hallopeau. gemini-3-flash-preview similarly included M161G Acrodermatitis continua and M1602 Arthritis mutilans, but also added specific parapsoriasis variants like ^ESCTGU266026 Guttate parapsoriasis, ^ESCTPA275907 Parapsoriasis lichenoides, and ^ESCTPA341403 Parapsoriasis en plaques. gpt-5.2 was the most precise, only including the irrelevant code ^ESCTPS666367 Generalised psoriasis due to a character mismatch with the target code. |
| Vascular dementia | Claude-sonnet-4 and GPT-5.2 primarily included optional codes defined in the criterion, such as ^ESCT1237065 **MVAD - Mixed vascular Alzheimer dementia** and ^ESCT1169441 **Subcortical dementia**. Claude-sonnet-4 also added Eu011-1 **[X]Predominantly cortical dementia** and Eu013 **Mixed cortical and subcortical vascular dementia**. Gemini-3-flash included the widest variety of irrelevant codes, incorporating optional items like ^ESCT1237064 **CVAD - Cortical vascular Alzheimer dementia**, ^ESCT1237066 **SVAD - Subcortical vascular Alzheimer dementia**, and ^ESCTMI802420 **Vascular dementia**. Unlike the other models, Gemini-3-flash also introduced codes entirely absent from the target list, specifically ^ESCTMU501126 **Multi-infarct state** and ^ESCTWH786117 **White matter disorder with CADASIL**. |
| Wrist fracture | Across several epochs, all models included irrelevant torus fracture codes not present in the gold standard. Claude-Sonnet-4-6 frequently included EMISR4QTO2 **Torus fracture of radius**. Gemini-3-Flash-Preview and GPT-5.2 both expanded on this by including ^ESCTTO651725 **Torus fracture of radius**, while Gemini-3-Flash-Preview also included ^ESCTCL710664 **Closed torus fracture of radius**. |
